# Causes of Neonatal Mortality in the European Region: A WHO-based analysis and Systematic Review

**DOI:** 10.1101/2025.07.01.25330618

**Authors:** Elizabeth E. la Cour, Veronica N. E. Malange, Tasnim Mohaissen, Michael Christiansen, Jesper Sune Brok, Christina Gade, Ulrik Lausten-Thomsen, Paula L. Hedley

**Affiliations:** Department for Congenital Disorders, Statens Serum Institut, Copenhagen, Denmark; Department of Biomedical Sciences, University of Copenhagen, Copenhagen, Denmark; Department of Epidemiology, College of Public Health, The University of Iowa, Iowa City, Iowa, USA; Department of Pediatric and Adolescent Medicine, Copenhagen University Hospital, Rigshospitalet, Copenhagen, Denmark; Department of Clinical Medicine, University of Copenhagen, Copenhagen, Denmark; Department of Clinical Pharmacology, Copenhagen University Hospital Bispebjerg and Frederiksberg, Copenhagen, Denmark; Department of Neonatology, University Hospital Rigshospitalet, Copenhagen, Denmark

**Keywords:** neonatal mortality, Europe, prematurity, congenital anomalies, ICD-PM, cause of death

## Abstract

**Purpose:** Despite considerable progress in reducing neonatal mortality, significant work remains. The regional differences in both the rates and causes of neonatal mortality, necessitates addressing this issue within a regional context. This study aimed to identify reported causes of neonatal death in the European Region and examine the cause-specific distribution of neonatal deaths over time.

**Methods:** We conducted a two-pronged analysis: (1) database analysis using WHO public datasets (2000–2021) for 28 countries in the European Region and (2) a systematic review and meta-analysis of pertinent records from various databases which were comprehensively reviewed against inclusion criteria up to 10^th^ August 2024, following PRISMA guidelines.

**Results:** The average neonatal mortality rate between 2000–2021 in WHO data was 2.63 per 1,000 live births, with a significant decline (-0.074 per year) across the European Region. Prematurity (41.2%) and congenital anomalies (28.9%) were the most common registered causes of neonatal death; no detailed etiological breakdown was available. The systematic review identified 41 eligible studies; 14 could be included in meta-analyses. Pooled estimates showed that congenital anomalies and prematurity each accounted for 30% of deaths. Among extremely preterm neonates, infections, respiratory and cardiovascular disorders were the most common reported causes of death.

**Conclusions:** NMRs across the European Region is declining (rapidly) with prematurity and congenital anomalies being leading causes of neonatal death. Current mortality reporting frameworks inadequately capture the diagnostic complexity of neonatal death causes. Adoption of detailed, standardized classification systems is critical to improving surveillance and data comparability, especially for preterm infants.

*What is Known:*

- Neonatal mortality rates have declined across Europe over recent decades.
- There are considerable regional disparities in both causes and rates of neonatal death.

*What is New:*

- Prematurity and congenital anomalies each account for ∼30% of neonatal deaths.
- In extremely preterm infants, infections and cardiorespiratory disorders predominate.
- Greater use of standardised classifications of neonatal death causes is urgently needed.

## 1 Introduction

The neonatal period accounts for the highest proportion of paediatric deaths worldwide [1; 2]. Consequently, reducing neonatal mortality is a global public health priority and an integral target under the United Nations’ ustainable Development Goals (SDGs), adopted in 2015 [3]. Through focused global and regional efforts, the neonatal mortality rate (NMR) has declined substantially from 37 deaths per 1,000 live births in 1990 to 17 in 2023, globally [1]. In Europe, this number has recently been reported to be extremely low, with NMR decreasing from 8 to 2 deaths per 1,000 live births over the same period [1].

Significant regional disparities persist in both NMRs and causes of death. Important causes include prematurity, intrapartum complications, congenital anomalies, and infections [1; 2]. In low-income settings, limited access to safe drinking water, sanitation, hygiene, and prenatal and neonatal care increases the risk of preventable deaths [1; 4]. Accordingly, it is essential to understand neonatal mortality patterns within their specific context.

Broader regional analyses allow for improved insight, with sufficient statistical power, into the causes of neonatal deaths, particularly when perinatal care is relatively uniform. However, the interpretation of neonatal mortality data is often hindered by non-specific cause-of-death classifications. For instance, “prematurity” is frequently recorded as a cause of death, despite being a risk factor rather than a definitive pathological mechanism [5].

To move beyond broad classifications and enable effective interventions, greater specificity in the categorization of neonatal deaths is essential. We aim to address that gap by systematically evaluating existing literature and publicly available databases to identify, classify, and synthesize the reported causes of neonatal death in the European region. A particular focus is placed on deaths occurring among extremely preterm infants, who represent a disproportionately vulnerable subgroup.

The primary objective of this systematic review is to improve understanding of the causes of neonatal death in the European Region by summarizing reports and databases on NMRs and causes.

## 2 Materials and Methods

### 2.1 Study Design and Registration

This study comprises two components: 1) a database analysis using publicly available data from World Health Organization (WHO) to examine causes of death and NMRs from 2000 to 2021. 2) a systematic review of literature encompassing studies published between 1^st^ January 2010 and 10^th^ August 2024 was conducted in accordance with the PRISMA 2020 guidelines [6]. The review protocol was developed following PRISMA-P guidance and is registered with PROSPERO (CRD42024571819).

### 2.2 Study Region

Analyses were restricted to Switzerland and all European Economic Area (EEA) member states as of 1 January 2010; hereafter referred to as the European Region.

### 2.3 Objectives and Outcomes

The primary objective was to identify specific reported causes of neonatal death in light of country-specific changes in NMRs. A secondary objective was to examine the causes of neonatal death among extremely preterm neonates (<28 weeks of gestation).

#### 2.3.1 Database Analysis Outcomes

The primary outcome was the cause-specific distribution of neonatal deaths evaluated using WHO classification standards. Additionally, the NMR per 1,000 live births, analysed by year and country.

#### 2.3.2 Systematic Review Outcomes

The main outcome was the proportion of neonatal deaths attributed to specific causes. These were reported separately for studies which included neonates of all gestational ages, and a secondary subset of studies that focused on extremely preterm neonates (defined above). Studies that examined other gestational age categories (i.e. term, very preterm, or moderate to late preterm) were excluded from further analysis.

### 2.4 Database Analysis

#### 2.4.1 Data Sources

The database analysis utilized WHO indicators for neonatal mortality [7], causes of death [8], and live birth statistics from United Nations Economic Commission for Europe (UNECE) [9].

#### 2.4.2 Statistical analysis

Descriptive statistics were used to summarise cause-specific mortality distributions and NMR trends. Linear regression models were fitted separately for each country, with NMR as the dependent variable and calendar year as an independent variable.

To identify regional patterns in cause-specific mortality, a two-step cluster analysis was conducted. First, hierarchical clustering using Ward’s minimum variance method was applied to standardised cause-specific mortality proportions and their corresponding linear trends across the study period. These variables were used to construct a distance matrix. The resulting dendrogram informed the optimal number of clusters. Next, K-means clustering was performed using the selected number of clusters to assign countries into distinct mortality profile groups. Thus, cluster definitions incorporated both the relative burden of specific causes of neonatal death and their temporal trends over the two-decade study period.

Statistical significance was evaluated using p-values, with values <0.05 considered statistically significant. All analyses were conducted in R version 4.4.2 (16).

### 2.5 Systematic Review and Meta-Analysis

#### 2.5.1 Search Strategy

A literature search was conducted on August 10, 2024, across PubMed, Embase, Web of Science, Google Scholar, Scopus, and the Cochrane Library (including CENTRAL). Prepublication servers (medRxiv, bioRxiv) were also searched. Additional references were identified through manual screening of bibliographies. Language translation, when required, was conducted using DocTranslator [10], and supported by language consultants as needed. A full search strategy is included in Supplementary Material.

#### 2.5.2 Eligibility Criteria

Eligible studies included cohort, case-control, cross-sectional, or interventional designs that reported on causes of neonatal death. Neonates were defined as per WHO criteria as infants under 28 days of age [11]; extremely preterm neonates were defined as those born before 28 weeks of gestation [12]. We defined neonatal death among preterm infants to be death within 28 days of the expected date of delivery.

The review was restricted to causes of neonatal death within the European Region. Furthermore, studies published before 2010 and studies published prior to 1984 were excluded.

#### 2.5.3 Data Management and Selection Process

Search results were managed in Rayyan [13]. After de-duplication, two reviewers (EELC and VNEM) independently screened titles and abstracts against predefined eligibility criteria. Articles were classified as “included”, “excluded”, or “maybe”. Full-text reviews were performed for all “included” and “maybe” articles. Disagreements were resolved through discussion or arbitration by a third reviewer (PLH). The selection process is outlined in the PRISMA flow diagram (Supplementary Figure 1) [6].

#### 2.5.4 Quality Assessment

The Newcastle-Ottawa Scale (NOS) [14] was used by two reviewers (EELC and PLH) to independently assess study quality. Discrepancies in scoring were resolved through consensus.

#### 2.5.5 Data Extraction

Two reviewers (EELC and PLH) independently extracted data from included studies, recording the author, publication year, study design, country, sample size, relevant findings, and NOS score. Studies reporting from overlapping datasets were cross-checked to prevent duplication.

#### 2.5.6 Data Synthesis and Classification

Causes of neonatal death were grouped into ICD-10 categories [15]: Perinatal conditions (P00–P96), Congenital malformations (Q00–Q99), and Other/unspecified (i.e. any cause not in chapters P or Q). Meta-analyses were performed only for categories P and Q. The “Other or unspecified” group was excluded due to heterogeneity by default.

In studies with sufficient diagnostic detail, causes of neonatal death were further stratified using the WHO application of ICD-10 to deaths during the perinatal period (ICD-PM) framework to enhance classification specificity [16] (Table 1). N3 (birth trauma) and N4 (intrapartum complications) were merged, as these categories were frequently reported together. N11 (unspecified causes) was excluded due to heterogeneity by default.

**Table 1:** Neonatal mortality categories defined by World Health Organisation’s application of the International Classification of Diseases, 10th Revision (ICD-10) to deaths during the perinatal period (ICD-PM). Table extracted from [16].

| Neonatal Death |  | ICD-10 codes |
| --- | --- | --- |
| ICD-PM:N1 | Congenital malformations, deformations and chromosomal abnormalities | Q00–Q99 |
| ICD-PM:N2 | Disorders related to fetal growth | P05, P08 |
| ICD-PM:N3 | Birth trauma | P10–P15 |
| ICD-PM:N4 | Complications of intrapartum events | P20, P21 |
| ICD-PM:N5 | Convulsions and disorders of cerebral status | P90, P91 |
| ICD-PM:N6 | Infection | P23, P35–P39 |
| ICD-PM:N7 | Respiratory and cardiovascular disorders | P22, P24–P29 |
| ICD-PM:N8 | Other neonatal conditions<br>including codes specific to the neonatal period from haemorrhagic and haematological disorders of fetus and newborn, transitory endocrine and metabolic disorders specific to fetus and newborn, digestive system disorders of fetus and newborn, conditions involving the integument and temperature regulation of fetus and newborn, other disorders originating in the perinatal period) | P50–P61, P70–P78, P80–P83, P92–P94 |
| ICD-PM:N9 | Low birth weight and prematurity | P07 |
| ICD-PM:N10 | Miscellaneous | P96.4 |
| ICD-PM:N11 | Neonatal death of unspecified cause | P96 |

#### 2.5.7 Statistical Analysis

Heterogeneity was quantified using I^2^ and Tau statistics (15). An I^2^ value >50% was considered indicative of substantial heterogeneity and >75% as considerable. Sensitivity analyses were performed, post-hoc, by iteratively excluding individual studies to assess the robustness of the pooled estimates.

Proportionate mortality for each cause of death was pooled across studies using a random-effects model due to high levels of heterogeneity. Meta-analyses were limited to studies reporting ICD-10 categories or ICD-PM categories with sufficient data. Categories with less than four studies or fewer than four deaths were excluded.

To explore publication bias, funnel plots were constructed for each cause-specific meta-analysis. Visual inspection of the funnel plots provided qualitative assessment of bias.

Statistical significance was determined using p-values, with values <0.05 considered statistically significant. All analyses were conducted in R version 4.4.2 (16).

## 3 Results

### 3.1 Database Analysis

#### 3.1.1 Causes of Neonatal Death and Neonatal Mortality Rates in the European Region

Prematurity was reported as the leading cause (41.2%), followed by congenital anomalies (28.9%) (Figure 1A). Both causes significantly declined over the study period (Figure 1B).

**Figure 1:**
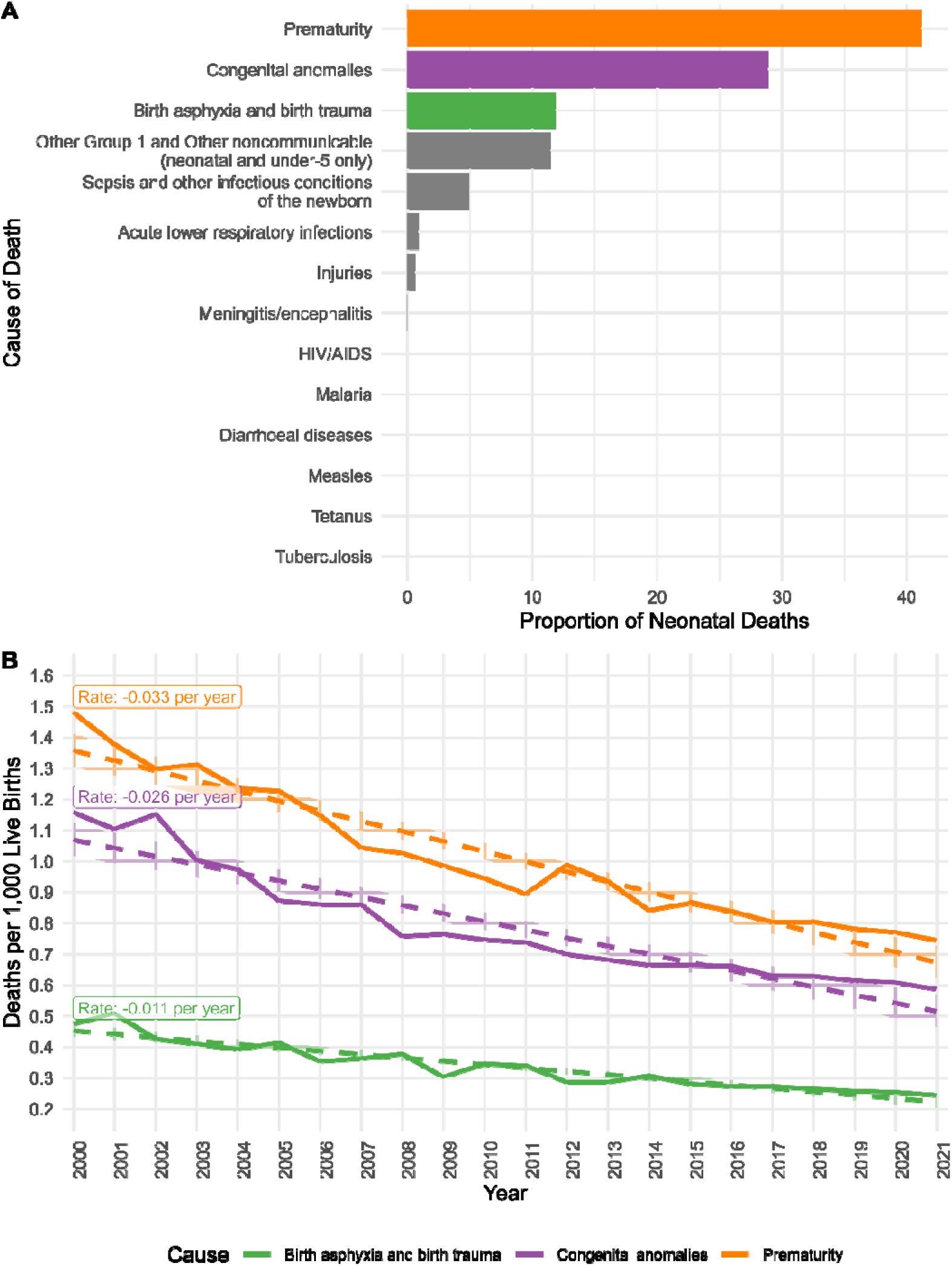
**A) Causes of Neonatal Death in the European Region, 2000–2021.** Proportionate mortality ratios for specific causes of neonatal death, based on combined data from WHO for the defined countries in the European region. **B) Top causes of Neonatal Death Over Time in the European Region, 2000–2021.** Trends in neonatal mortality rates per 1,000 live births for the three leading causes of death. Solid lines represent annual mortality rates. Dashed lines represent the linear regression models. Shaded areas show the 95%-confidence interval for each regression model. All causes showed significant decline between 2000 and 2021 (P < 0.001). The rate of decline for each cause is labelled.

Data on NMR from 2000–2021 were extracted from WHO databases for 28 European countries; Liechtenstein was excluded due to lack of data [7–9] (Supplementary Table 1 and Supplementary Figure 2). All countries showed statistically significant declining NMRs, with the exception of France (p = 0.264). The average NMR across the European region was 2.63 deaths per 1,000 live births, with a significant decline of -0.074 per year (p < 0.001) (Supplementary Table 1).

#### 3.1.2 Cluster Analysis

Hierarchical clustering analysis grouped the Baltic countries into one cluster, while the remaining countries formed a second cluster (Supplementary Figure 2 and Figure 2). Key differentiators included lower rates of death due to prematurity and higher rates of death due to birth asphyxia in the Baltic countries, as well as differing trends in leading causes of death over time (Figure 3).

**Figure 2:**
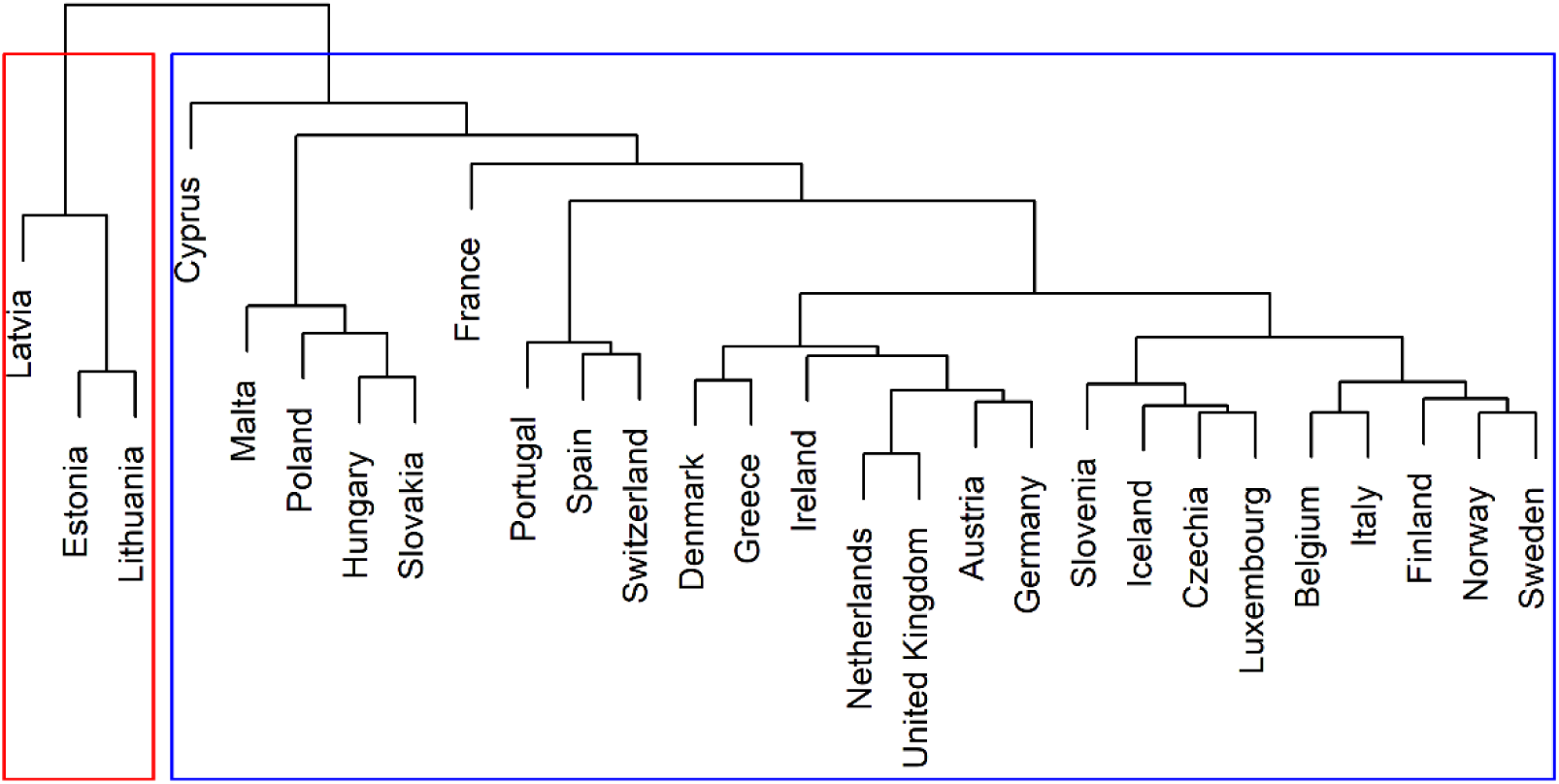
Hierarchical clustering of countries in the European Region based on neonatal mortality causes and trends. The dendrogram illustrates the results of hierarchical clustering using Ward’s minimum variance method applied to standardised cause-specific mortality proportions and their linear trends across the study period. Two main clusters are identified: A smaller cluster (red) with Latvia, Estonia and Lithuania, and a larger cluster (blue) with the remaining countries. This model illustrates regional similarities and differences in mortality rates and causes of death.

**Figure 3:**
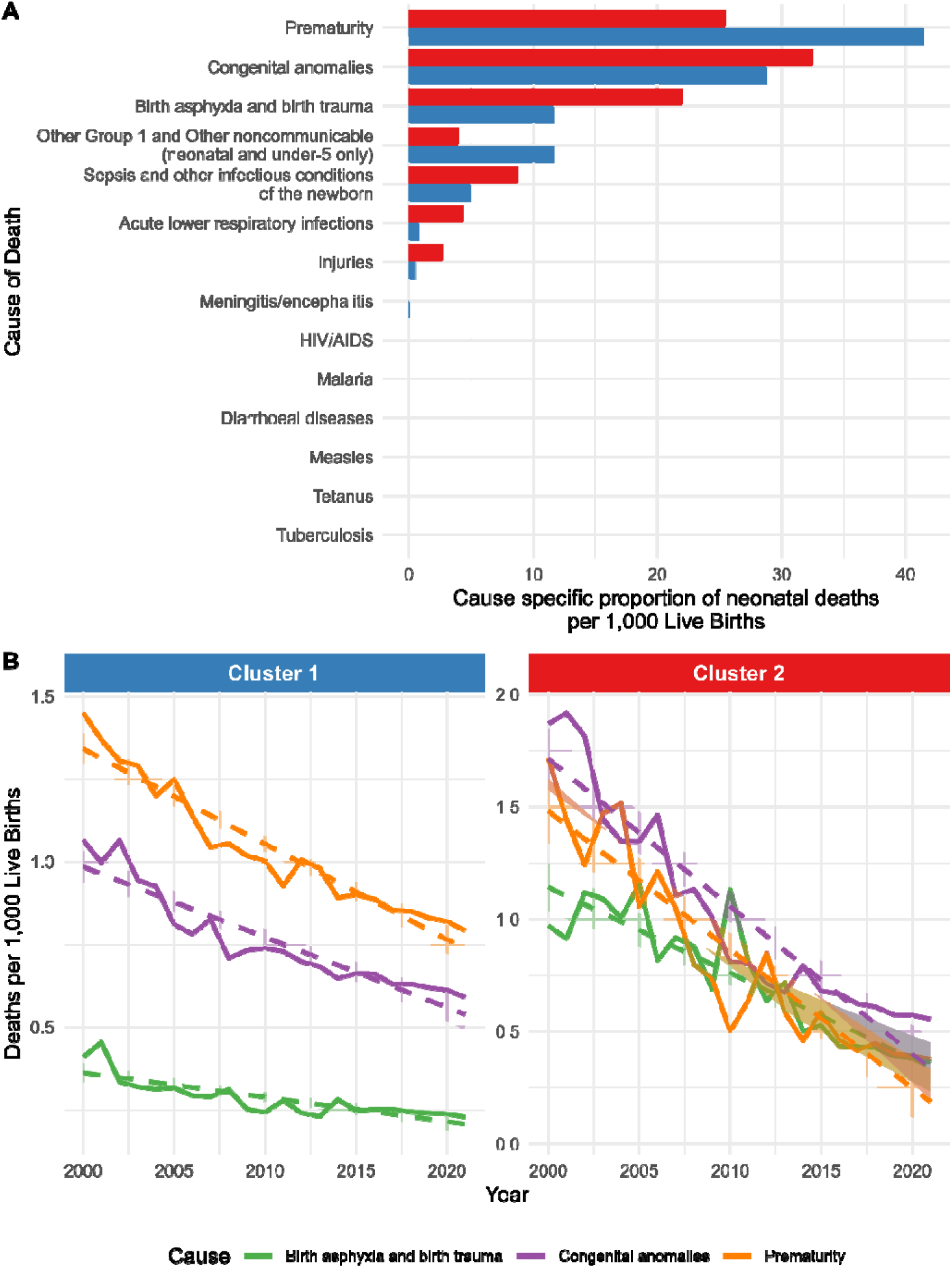
**A) Causes of Neonatal Death divided in clusters of European countries, 2000–2021.** Proportionate mortality ratios for specific causes of neonatal death, based on combined data from WHO for the defined clusters in the European region. Cluster 1 (blue): all countries with the exception of those in cluster 2 (red): Latvia, Estonia and Lithuania. **B) Top causes of Neonatal Death Over Time divided in clusters of European countries, 2000–2021.** Trends in neonatal mortality rates per 1,000 live births for the three leading causes of death. Solid lines represent annual mortality rates. Dashed lines represent the linear regression models. Shaded areas show the 95%-confidence interval for each regression model. All causes showed significant decline between 2000 and 2021 (P < 0.001). Cluster 1: all countries with the exception of those in cluster 2: Latvia, Estonia and Lithuania.

### 3.2 Systematic Review and Meta-Analysis

#### 3.2.1 Study Selection and Characteristics

Out of 4,644 identified articles, 1,320 duplicates were removed. After title and abstract screening of the remaining 3,324 articles, 601 underwent full-text review. Forty articles and two WHO databases met inclusion criteria, with one additional article included through reference list screening (Supplementary Figure 1).

The 41 included studies covered 17 countries. The most frequently represented countries were the United Kingdom (n=14), Denmark (n=4), and Poland (n=4). No eligible studies were identified from the Baltic countries. Sample sizes ranged from 71 to over 6 million neonates. Neonatal death cohorts ranged from 4 to 13,077 cases. Reporting on multiple births was inconsistent, with only 39% of studies specifying whether the cases involved singleton of multiple births. Four studies were in non-English European languages and translated using an online tool [10]. An overview of the included studies is provided in Supplementary Table 2

The study with the highest neonatal death incidence was Gatt et al., analysing Maltese births from 1994–2013 (n=84,821) [17].

#### 3.2.2 Classification and Quality of Studies

Death cause classification methods varied and included ICD-PM [16], CODAC [18], Wigglesworth [19], INCODE- based methods [20], and the system used by Patel et al. [21]. Six studies focused on NICU deaths [12; 22-26], while others such as Basu et al. [20], Troszynski et al. [27], and Miranda et al. [28] included only early neonatal deaths (<7 days). Some studies offered detailed temporal or age-stratified classifications, e.g., Williams et al. [29], and Pantou et al. [30].

Seven studies used forensic assessments and autopsy-based classification [31–37]. Liu et al. and Smith et al. [38; 39] used the Global Burden of Disease dataset. Thirty-one studies were cross-sectional and 10 were cohort studies; NOS scores ranged from 2 to 9 stars [14] (Supplementary Table 2).

#### 3.2.3 Heterogeneity and Publication Bias

Substantial heterogeneity was observed across all pooled analyses (I^2^ > 50%). Funnel plot inspection suggested possible publication bias in most analyses. Post-hoc sensitivity analyses excluding individual study outliers had minimal effect on I^2^ or Tau^2^.

#### 3.2.4 Meta-Analysis: Neonates of All Gestational Ages

Fourteen studies were eligible for meta-analysis (Table 2 and Table 3), yielding eleven general neonatal cohorts. Studies reporting only data on term neonates (e.g., Ali et al. [12], and Basu et al. [20]) were excluded, as was an unmatched preterm sub-cohort from Allanson et al. [5]. Meta-analysis results showed that 55% of neonatal deaths were attributed to ICD-10 perinatal conditions (95% CI: 46–64%) and 30% were attributed to congenital malformations (95% CI: 21–41%). Among ICD-PM categories, the groups with highest mortality were N1 (congenital anomalies) at 30% (95% CI: 18–47%) and N9 (low birth weight/prematurity), also at 30% (95% CI: 17–46%). The other ICD-PM categories each accounted for <11% of deaths. Due to insufficient data, no analysis was conducted for N2, N5, N8, or N10 (defined in Table 1).

**Table 2:** Data extraction sheet for meta-analyses. ICD-10. Data on neonatal mortality causes were classified into ICD-10 categories. NOS scores are indicated below the study name. n.d. = no data. a = indicates ly neonatal death cohorts. b = indicates the older cohort in a study which compares two time periods.

| Study<br>(NOS score) | Period | Duration<br>months | Country | Level | Singleton/<br>Multiples | Included GA | GA group | Death period | Number of<br>deaths | Perinatal<br>conditions<br>(P00-P96) | Congenital<br>malformations<br>(Q00-Q99) | Other or<br>unspecified |
| --- | --- | --- | --- | --- | --- | --- | --- | --- | --- | --- | --- | --- |
| Allanson et al. (2016)<br>(00000000) | 1997- 2010 | 156 | UK | County | Both | > 20+0 | all | Neonatal<br>deaths | 4204 | 1614 | 1152 | 1438 |
| Best et al. (2019)<br>(00000000) | 2014-2015 | 12 | UK | National | Singletons | > 24+0 | all | Neonatal<br>deaths | 2345 | 1258 | 774 | 313 |
| Carty et al. (2023)<br>(0000000000) | 2005-2014 | 108 | England | National | Singletons | > 22+0 | all | Neonatal<br>deaths | 13077 | 8398 | 4070 | 609 |
| Costeloe et al. (2012)<sup>b</sup><br>(00000000) | 1 March- 31<br>December 1995 | 10 | England | National | n.d. | 22+0 - 25+6 | extremely<br>preterm | NICU deaths | 400 | 176 | 6 | 218 |
| Costeloe et al. (2012)<br>(00000000) | 2006 | 12 | England | National | n.d. | 22+0 - 26+6 | extremely<br>preterm | NICU deaths | 522 | 356 | 4 | 162 |
| Miranda et al. (2020)<sup>a</sup><br>(00000000) | 2008- 2017 | 108 | Portugal | NICU | n.d. | > 23+0 | all | Early Neonatal<br>deaths | 76 | 2 | 56 | 18 |
| Nohr et al. (2021)<br>(0000000000) | 1996-2002 | 72 | Denmark | National | Singletons | > 22+0 | all | Neonatal<br>deaths | 226 | 81 | 91 | 54 |
| Pantou et al. (2010)<sup>a</sup><br>(00000000) | 1996-2004 | 96 | Greece | Regional<br>& NICU | n.d. | > 22+0 | all | Early Neonatal<br>deaths | 41 | 33 | 5 | 3 |
| Pantou et al. (2010)<br>(00000000) | 1996-2004 | 96 | Greece | Regional<br>& NICU | n.d. | > 22+0 | all | Neonatal death | 103 | 53 | 11 | 39 |
| Stensvold et al. (2017)<br>(0000000000) | 2013-2014 | 12 | Norway | National | both | 22+0 - 26+6 | extremely<br>preterm | NICU deaths | 66 | 32 | 2 | 32 |
| Tambe et al (2015)<br>(00000000) | 2006-2008 | 24 | UK | National | n.d. | n.d. | all | Neonatal<br>deaths | 8960 | 6817 | 1575 | 568 |
| Tambe et al (2015)<br>(00000000) | 2006-2008 | 24 | Sweden | National | n.d. | n.d. | all | Neonatal<br>deaths | 574 | 307 | 158 | 109 |
| Troszynski et al. (2011)<sup>a</sup><br>(00000000) | 2007-2009 | 24 | Poland | Regional | n.d. | n.d. | all | Early Neonatal<br>deaths | 1412 | 683 | 435 | 294 |
| van Beek et al. (2021)<sup>b</sup><br>(0000000000) | 2007-2009 | 24 | Netherlands | National | Both | 24+0 - 26+6 | extremely<br>preterm | NICU deaths | 203 | 131 | 1 | 71 |
| van Beek et al. (2021)<br>(0000000000) | 2011-2017 | 72 | Netherlands | National | Both | 24+0 - 26+6 | extremely<br>preterm | NICU deaths | 603 | 441 | 7 | 155 |
| Verhagen et al. (2010)<br>(00000000) | 2005- 2006 | 12 | Netherlands | NICU | n.d. | > 22+6 | all | NICU deaths | 52 | 26 | 26 | 0 |

**Table 3:**
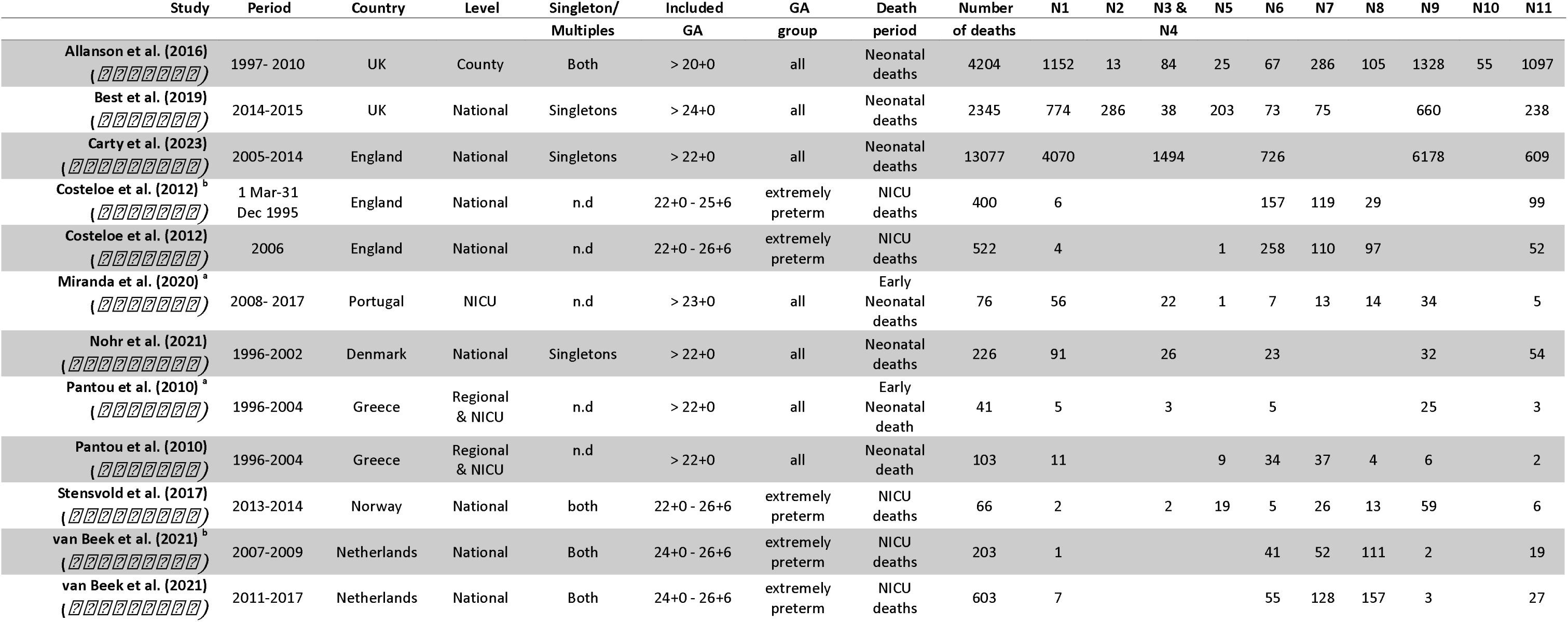
Data extraction sheet for meta-analyses. ICD-PM. Data on neonatal mortality causes were classified into ICD-PM categories, described in Table 1. NOS scores are indicated below the study name. n.d. = data. a = indicates early neonatal death cohorts. b = indicates the older cohort in a study which compares two time periods.

#### 3.2.5 Meta-Analysis: Extremely Preterm Neonates

Five cohorts from three studies focused on extremely preterm neonates (Table 2 and Table 3). Meta-analyses showed that 61% of deaths were due to ICD-10 perinatal conditions (95% CI: 50–70%) and ICD-PM categories N6 (Infection), N7 (Respiratory and cardiovascular disorders), and N8 (Other neonatal conditions) each accounted for 21–26% of deaths.

## 4 Discussion

This study aimed to explore the causes of neonatal deaths in the European Region and examine the cause- specific distribution of neonatal deaths over time, with a particular focus on extremely preterm neonates. Based on the WHO data [7], the leading causes were reported to be prematurity (41.2%) and congenital anomalies (28.9%). All the major causes of death were declining over the study period.

The WHO database is a key global resource for neonatal mortality surveillance. However, its global structure may not suit the specific analytical needs of high-income regions such as Europe. Several cause-of-death categories (e.g., HIV/AIDS, malaria, diarrhoeal diseases, tetanus, tuberculosis), rarely reported in European countries due to high levels of hygiene and universal healthcare access [4; 12], may obscure regionally relevant conditions [2].

The systematic review identified 41 relevant studies from an initial pool of 4,644 citations. Many were excluded due to broad outcome measures, such as “infant mortality” or “perinatal death”, that precluded meaningful neonatal-specific analysis. NICU-based death reports were retained, acknowledging that many preterm infants may be chronologically older than 28 days at death but still fall within the neonatal period [22; 23]. A major limitation of the existing literature is the inconsistency in classification systems. Studies often used vague or ambiguous categories such as “neonatal” or “prematurity” without further detail. For instance, Allanson et al. reported that 77.3% of neonatal deaths in the UK occurred in preterm infants, but without more precise coding, actionable insights remain limited [5]. The absence of standardized classification frameworks creates heterogeneity across studies and countries. Variations in live birth definitions, abortion policies, and prenatal diagnostics may influence how deaths are registered and interpreted [40]. Without standardized approaches, producing reliable cross-country comparisons and pooled estimates remains a challenge.

This review adhered to PRISMA guidelines [6] and implemented a comprehensive literature search. Dual independent reviewers performed screening, data extraction, and quality appraisal using the NOS scale [14]. However, methodological challenges were encountered. To standardize data, all study-reported causes were recoded into ICD-PM categories [16], which may have introduced misclassification despite best efforts. Given the low incidence of neonatal death in the included countries, many studies were based on small sample sizes. While exclusion based on study size was avoided to minimize selection bias, smaller cohorts contributed to substantial heterogeneity in the meta-analyses.

A significant concern is the dominant use of the term “prematurity” as a cause of death. As a classification, “prematurity” lacks specificity, analogous to using “old age” in adult death certificates and fails to capture the underlying pathologies that lead to neonatal death in preterm infants. This is particularly relevant for extremely preterm infants, among whom twin pregnancies are overrepresented (∼10%). The distinct physiological profiles of these infants and elevated risk of complications may influence the types and frequencies of underlying causes of death recorded. Given that most neonatal deaths occur among preterm infants [41], this lack of detail restricts our understanding and limits the clinical utility of the data for guiding interventions.

National policies around abortion and neonatal viability thresholds affect reported mortality distributions [39]. Differences in the proportion of neonatal deaths attributed to congenital malformations and prematurity across countries may, in part, reflect underlying disparities in prenatal care, pregnancy termination practices, and obstetric decision-making. Severe congenital anomalies are often detectable through routine antenatal screening programmes [42] and in countries where pregnancy termination is both legally permitted and culturally accepted, such cases may be electively terminated, thereby reducing the number of neonates born with fatal anomalies. Conversely, in settings where abortion is legally restricted or socially discouraged, these pregnancies may continue to term, resulting in a higher proportion of neonatal deaths from congenital malformations. Similarly, the timing of preterm birth is not solely determined by spontaneous physiological processes but is also shaped by clinical practice. National obstetric guidelines influence when and under what conditions high-risk pregnancies, such as those complicated with foetal growth restriction, pre-eclampsia or other maternal-foetal indications, are electively delivered prematurely. Variability in these guidelines and the availability of perinatal resources likely contributes to inter-country differences in the distribution and timing of preterm births, and subsequently, to the observed patterns of neonatal mortality.

Improved specificity in neonatal death reporting is necessary to enable targeted prevention strategies. Future studies should adopt standardized, detailed classification systems and explicitly define inclusion criteria for neonatal age brackets (e.g., early vs. late neonatal death), as well as multi-fetal vs singleton pregnancies. Vague umbrella categories like “preterm birth” or “other” should be avoided. More frequent use of autopsies could also substantially improve diagnostic accuracy. For example, Liebrechts-Akkerman et al. demonstrated that autopsy findings provided critical insight into causes of death not captured by standard certification [34]. Yet, autopsy rates have declined in countries such as Denmark [43], and re-expanding their use in clinical practice will require addressing ethical, logistical, and financial barriers. Establishing standardized mortality classifications across Europe would strengthen the validity of national datasets and improve the reliability of cross-border comparisons. Harmonized registration practices, integrated with robust classification systems such as ICD-PM, would enhance the depth of epidemiological analysis and inform health policy.

## 5 Conclusions

This study demonstrated that prematurity and congenital anomalies were the leading causes of neonatal death in a frame of decreasing NMRs across the European Region between 2000 and 2021. However, broad cause of death classifications reflects the limited granularity of current data sources, particularly the WHO database, which lacks diagnostic detail suited to high-income settings. The systematic review of the literature revealed considerable heterogeneity in classification methods and outcome reporting across studies. This variability underscores the urgent need for standardized, detailed frameworks, such as ICD-PM, for neonatal mortality reporting.

A consistent and granular approach to classifying neonatal deaths, particularly among preterm neonates, is essential to advance the accuracy of epidemiological data, enable meaningful international comparisons, and guide more effective clinical and public health interventions.

## Data Availability

All data produced in the present work are contained in the manuscript

## List of Abbreviations

EEA: European Economic Area
ICD-10: International Classification of Diseases, 10th Revision
ICD-PM: The WHO application of ICD-10 to deaths during the perinatal period
LB: live births
Lm coeff: linear regression coefficient
NMR: neonatal mortality rate
NOS: Newcastle-Ottawa Scale
SD: standard deviation
SDGs: United Nations’ Sustainable Development Goals
UNECE: United Nations Economic Commission for Europe
WHO: World Health Organization

## Statements and Declarations

### Author Contributions

Elizabeth la Cour, Michael Christiansen, Ulrik Lausten-Thomsen, and Paula L. Hedley contributed to the study conception and design. Material preparation, data collection and analysis were performed by Elizabeth la Cour, Veronica N. E. Malange and Paula L. Hedley. The first draft of the manuscript was written by Elizabeth la Cour and all authors commented on previous versions of the manuscript. All authors read and approved the final manuscript.

