## Supplementary Materials for "Causes of Neonatal Mortality in the European Region: A WHO-based analysis and Systematic Review"

### Supplementary Material: Search Strategy

1. Pubmed

2. Embase

3. Web of Science core collection

4. Google Scholar

5. Scopus

6. Cochrane Library including CENTRAL

7. Prepublication server medRXiv and bioRXiv

#### PubMed

**Supplementary Table 1:** List of search terms in PubMed and number of search results form a search performed

on the 10th of August 2024.

| Aspect 1 |  | Aspect 2 |
| --- | --- | --- |
| "infant, newborn"[MeSH Terms] | AND | "Death"[MeSH Terms] |
| AND |  | OR |
| "Neonat*"[Title/Abstract] |  | "Infant Death"[MeSH Terms] |
| OR |  | OR |
| "Newborn*"[Title/Abstract] |  | "Perinatal Death"[MeSH Terms] |
|  |  | OR |
|  |  | "Cause of Death"[MeSH Terms] |
|  |  | OR |
|  |  | "Infant Mortality"[MeSH Terms] |
|  |  | AND |
|  |  | "Death Cause*"[Title/Abstract] |
|  |  | OR |
|  |  | "Cause* of Death"[Title/Abstract] |
|  |  | OR |
|  |  | "determinant* of mortalit*"[Title/Abstract] |
|  |  | OR |
|  |  | "Mortalit* Determinant*"[Title/Abstract] |
|  |  | OR |
|  |  | “Neonat* Outcome*”[Title/Abstract] |
|  |  | OR |
|  |  | “Infant* outcome*”[Title/Abstract] |

1712 results with a date limit of 1^st^ January 2010. Search made on the 10th August 2024

("infant, newborn"[MeSH Terms] AND ("Neonat*"[Title/Abstract] OR "Newborn*"[Title/Abstract])) AND

(("Death"[MeSH Terms] OR "Infant Death"[MeSH Terms] OR "Perinatal Death"[MeSH Terms] OR "Cause of

Death"[MeSH Terms] OR "Infant Mortality"[MeSH Terms]) AND ("Death Cause*"[Title/Abstract] OR "Cause* of

Death"[Title/Abstract] OR "determinant* of mortalit*"[Title/Abstract] OR "Mortalit* Determinant*"[Title/Abstract]

OR “Neonat* Outcome*”[Title/Abstract] OR “Infant* outcome*”[Title/Abstract]))

#### Embase

**Supplementary Table 2:** List of search terms in Embase and number of search results form a preliminary search

performed on the 10th of august 2024.

| Aspect 1 |  | Aspect 2 |  | Aspect 3 |
| --- | --- | --- | --- | --- |
| Infant (subject heading) | AND | Neonat* (title) | AND | "Death Cause*"(Title/Abstract) |
| OR |  | OR |  | OR |
| Baby (subject heading) |  | Newborn* (title) |  | "Cause* of Death"(Title/Abstract) |
| OR |  | OR |  | OR |
| Newborn (subject heading) |  | Infant* (title) |  | "determinant* of mortalit*"(Title/Abstract) |
|  |  |  |  | OR |
| AND |  | AND |  | "Mortalit* Determinant*"(Title/Abstract) |
|  |  |  |  | OR |
| “Cause of death” (subject heading) |  | Death*(title) |  | Outcome* (abstract) |
|  |  | OR |  | OR |
| OR |  | Mortalit*(title) |  | Survival* (abstract) |
|  |  | OR |  |  |
| “infant mortality” (subject heading) |  | Outcome*(title) |  |  |
| OR |  | OR |  |  |
| “newborn death” (subject heading) |  | Survival*(title) |  |  |
| AND |  |  |  |  |
| Cause* (title/abstract) |  |  |  |  |

1183 results with a date limit of 1^st^ January 2010. Search made on the 10th August 2024

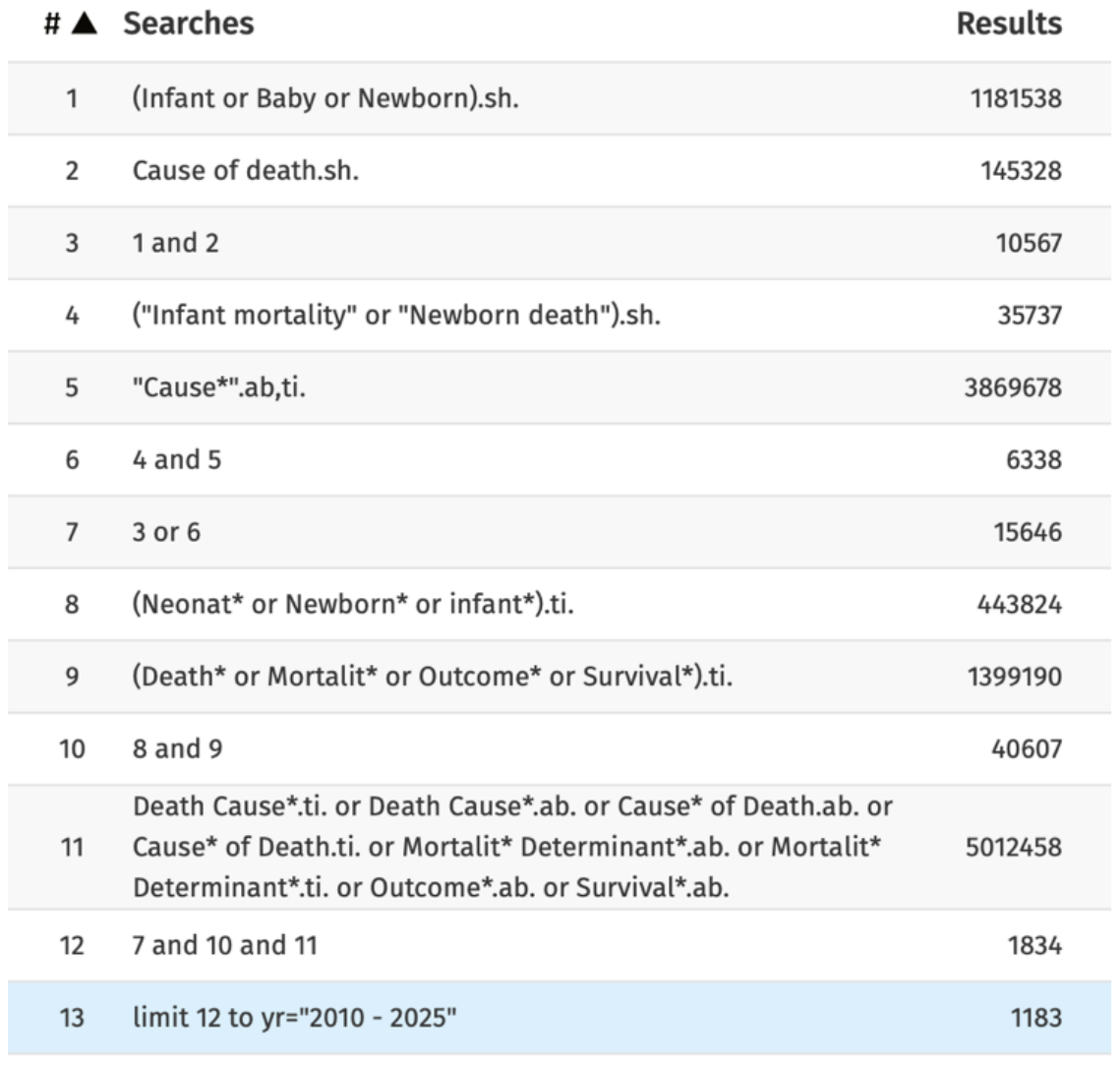

#### Web of Science core collection

Search made in “title”, date limit of 1^st^ January 2010.

(newborn OR newborns OR neonate OR neonates OR neonatal) AND (Death OR Deaths OR Mortalities OR mortality) AND (Cause OR Causes OR determinant OR determinants OR outcome OR outcomes)

Search made on the 10th August 2024 = 427 results

#### Google Scholar

(newborn OR newborns OR neonate OR neonates OR neonatal) AND (Death OR Deaths OR mortalities OR mortality) AND (Cause OR Causes OR determinant OR determinants OR outcome OR outcomes)

Search made on the 10th August 2024. In the articles “title”, date limit of 1^st^ January 2010.

Newborn AND Death AND Cause = 13 results

Newborn AND death AND causes = 9

Newborn AND Death AND determinant = 0

Newborn AND death AND determinants = 0

Newborn AND Death AND outcome = 3

Newborn AND death AND outcomes = 2

Newborn AND Mortality AND Cause = 4

Newborn AND mortality AND causes = 11

Newborn AND mortality AND determinant = 0

Newborn AND mortality AND determinants = 9

Newborn AND mortality AND outcome = 7

Newborn AND mortality AND outcomes = 4

Newborns AND Mortality AND Cause = 2

Newborns AND mortality AND causes = 8

Newborns AND mortality AND determinant = 0

Newborns AND mortality AND determinants = 11

Newborns AND mortality AND outcome = 7

Newborns AND mortality AND outcomes = 2

Neonate AND Death AND Cause = 3

Neonate AND death AND causes = 0

Neonate AND Death AND determinant = 0

Neonate AND death AND determinants = 0

Neonate AND Death AND outcome = 0

Neonate AND death AND outcomes = 0

Neonate AND Mortality AND Cause = 4

Neonate AND mortality AND causes = 0

Neonate AND mortality AND determinant = 0

Neonate AND mortality AND determinants = 0

Neonate AND mortality AND outcome = 0

Neonate AND mortality AND outcomes = 0

Neonates AND Mortality AND Cause = 9

Neonates AND mortality AND causes = 13

Neonates AND mortality AND determinant = 1

Neonates AND mortality AND determinants = 7

Neonates AND mortality AND outcome = 13

Neonates AND mortality AND outcomes = 19

Neonatal AND Death AND Cause = 43

Neonatal AND death AND causes = 112

Neonatal AND Death AND determinant = 1

Neonatal AND death AND determinants = 10

Neonatal AND Death AND outcome = 16

Neonatal AND death AND outcomes = 11

Neonatal AND Mortality AND Cause = 64

Neonatal AND mortality AND causes = 151

Neonatal AND mortality AND determinant = 10

Neonatal AND mortality AND determinants = 170

Neonatal AND mortality AND outcome = 32

Neonatal AND mortality AND outcomes = 91

Newborn AND Deaths AND Cause = 2

Newborn AND deaths AND causes = 6

Newborn AND Deaths AND determinant = 0

Newborn AND deaths AND determinants = 0

Newborn AND Deaths AND outcome = 1

Newborn AND deaths AND outcomes = 2

Newborns AND Deaths AND Cause = 2

Newborns AND deaths AND causes = 1

Newborns AND Deaths AND determinant = 0

Newborns AND deaths AND determinants = 1

Newborns AND Deaths AND outcome = 0

Newborns AND deaths AND outcomes = 0

Neonate AND Deaths AND Cause = 0

Neonate AND deaths AND causes = 1

Neonate AND Deaths AND determinant = 0

Neonate AND deaths AND determinants = 0

Neonate AND Deaths AND outcome = 0

Neonate AND deaths AND outcomes = 0

Neonates AND Deaths AND Cause = 1

Neonates AND deaths AND causes = 4

Neonates AND Deaths AND determinant = 0

Neonates AND deaths AND determinants = 0

Neonates AND Deaths AND outcome = 0

Neonates AND deaths AND outcomes = 0

Neonatal AND Deaths AND Cause = 24

Neonatal AND deaths AND causes = 58

Neonatal AND Deaths AND determinant = 0

Neonatal AND deaths AND determinants = 15

Neonatal AND Deaths AND outcome = 1

Neonatal AND deaths AND outcomes = 8

Newborn AND Mortalities AND Cause = 0

Newborn AND mortalities AND causes = 0

Newborn AND mortalities AND determinant = 0

Newborn AND mortalities AND determinants = 1

Newborn AND mortalities AND outcome = 0

Newborn AND mortalities AND outcomes = 0

Newborns AND mortalities AND Cause = 0

Newborns AND mortalities AND causes = 0

Newborns AND mortalities AND determinant = 0

Newborns AND mortalities AND determinants = 0

Newborns AND mortalities AND outcome = 0

Newborns AND mortalities AND outcomes = 0

Neonate AND mortalities AND Cause = 0

Neonate AND mortalities AND causes = 0

Neonate AND mortalities AND determinant = 0

Neonate AND mortalities AND determinants = 0

Neonate AND mortalities AND outcome = 0

Neonate AND mortalities AND outcomes = 0

Neonates AND mortalities AND Cause = 0

Neonates AND mortalities AND causes = 0

Neonates AND mortalities AND determinant = 0

Neonates AND mortalities AND determinants = 0

Neonates AND mortalities AND outcome = 0

Neonates AND mortalities AND outcomes = 0

Neonatal AND mortalities AND Cause = 0

Neonatal AND mortalities AND causes = 1

Neonatal AND mortalities AND determinant = 0

Neonatal AND mortalities AND determinants = 4

Neonatal AND mortalities AND outcome = 1

Neonatal AND mortalities AND outcomes = 1

In total: 869 results

#### Scopus

Search made in “article title”, date limit of 1^st^ January 2010.

(newborn OR newborns OR neonate OR neonates OR neonatal) AND (Death OR Deaths OR mortalities OR mortality) AND (Cause OR Causes OR determinant OR determinants OR outcome OR outcomes)

Search made on the 10th August 2024 = 423 results

#### Cochrane Library including CENTRAL

Search made in “record title”, date limit of 1^st^ January 2010.

(newborn OR newborns OR neonate OR neonates OR neonatal) AND (Death OR Deaths OR mortalities OR mortality)

AND (Cause OR Causes OR determinant OR determinants OR outcome OR outcomes)

Search made on the 10th August 2024 = 23 results

#### Prepublication server medRXiv and bioRXiv

(newborn OR newborns OR neonate OR neonates OR neonatal) AND (Death OR Deaths OR mortalities OR mortality) AND (Cause OR Causes OR determinant OR determinants OR outcome OR outcomes) Only prepublications from within 18 months were searched, to avoid finding already published articles.

Search made on the 10th August 2024. In the articles title, date limit of 1^st^ January 2010.

Newborn AND Death AND Cause = 0

Newborn AND death AND causes = 0

Newborn AND Death AND determinant = 0

Newborn AND death AND determinants = 0

Newborn AND Death AND outcome = 0

Newborn AND death AND outcomes = 0

Newborn AND Mortality AND Cause = 0

Newborn AND mortality AND causes = 0

Newborn AND mortality AND determinant = 0

Newborn AND mortality AND determinants = 0

Newborn AND mortality AND outcome = 0

Newborn AND mortality AND outcomes = 0

Newborns AND Mortality AND Cause = 0

Newborns AND mortality AND causes = 0

Newborns AND mortality AND determinant = 0

Newborns AND mortality AND determinants = 0

Newborns AND mortality AND outcome = 0

Newborns AND mortality AND outcomes = 0

Neonate AND Death AND Cause = 0

Neonate AND death AND causes = 0

Neonate AND Death AND determinant = 0

Neonate AND death AND determinants = 0

Neonate AND Death AND outcome = 0

Neonate AND death AND outcomes = 0

Neonate AND Mortality AND Cause = 0

Neonate AND mortality AND causes = 0

Neonate AND mortality AND determinant = 0

Neonate AND mortality AND determinants = 0

Neonate AND mortality AND outcome = 0

Neonate AND mortality AND outcomes = 0

Neonates AND Mortality AND Cause = 0

Neonates AND mortality AND causes = 0

Neonates AND mortality AND determinant = 0

Neonates AND mortality AND determinants = 0

Neonates AND mortality AND outcome = 0

Neonates AND mortality AND outcomes = 0

Neonatal AND Death AND Cause = 0

Neonatal AND death AND causes = 1

Neonatal AND Death AND determinant = 0

Neonatal AND death AND determinants = 1

Neonatal AND Death AND outcome = 0

Neonatal AND death AND outcomes = 0

Neonatal AND Mortality AND Cause = 1

Neonatal AND mortality AND causes = 1

Neonatal AND mortality AND determinant = 0

Neonatal AND mortality AND determinants = 2

Neonatal AND mortality AND outcome = 0

Neonatal AND mortality AND outcomes = 2

Newborn AND Deaths AND Cause = 0

Newborn AND deaths AND causes = 0

Newborn AND Deaths AND determinant = 0

Newborn AND deaths AND determinants = 0

Newborn AND Deaths AND outcome = 0

Newborn AND deaths AND outcomes = 0

Newborns AND Deaths AND Cause = 0

Newborns AND deaths AND causes = 0

Newborns AND Deaths AND determinant = 0

Newborns AND deaths AND determinants = 0

Newborns AND Deaths AND outcome = 0

Newborns AND deaths AND outcomes = 0

Neonate AND Deaths AND Cause = 0

Neonate AND deaths AND causes = 0

Neonate AND Deaths AND determinant = 0

Neonate AND deaths AND determinants = 0

Neonate AND Deaths AND outcome = 0

Neonate AND deaths AND outcomes = 0

Neonates AND Deaths AND Cause = 0

Neonates AND deaths AND causes = 0

Neonates AND Deaths AND determinant = 0

Neonates AND deaths AND determinants = 0

Neonates AND Deaths AND outcome = 0

Neonates AND deaths AND outcomes = 0

Neonatal AND Deaths AND Cause = 0

Neonatal AND deaths AND causes = 0

Neonatal AND Deaths AND determinant = 0

Neonatal AND deaths AND determinants = 0

Neonatal AND Deaths AND outcome = 0

Neonatal AND deaths AND outcomes = 0

Newborn AND Mortalities AND Cause = 0

Newborn AND mortalities AND causes = 0

Newborn AND mortalities AND determinant = 0

Newborn AND mortalities AND determinants = 0

Newborn AND mortalities AND outcome = 0

Newborn AND mortalities AND outcomes = 0

Newborns AND mortalities AND Cause = 0

Newborns AND mortalities AND causes = 0

Newborns AND mortalities AND determinant = 0

Newborns AND mortalities AND determinants = 0

Newborns AND mortalities AND outcome = 0

Newborns AND mortalities AND outcomes = 0

Neonate AND mortalities AND Cause = 0

Neonate AND mortalities AND causes = 0

Neonate AND mortalities AND determinant = 0

Neonate AND mortalities AND determinants = 0

Neonate AND mortalities AND outcome = 0

Neonate AND mortalities AND outcomes = 0

Neonates AND mortalities AND Cause = 0

Neonates AND mortalities AND causes = 0

Neonates AND mortalities AND determinant = 0

Neonates AND mortalities AND determinants = 0

Neonates AND mortalities AND outcome = 0

Neonates AND mortalities AND outcomes = 0

Neonatal AND mortalities AND Cause = 0

Neonatal AND mortalities AND causes = 0

Neonatal AND mortalities AND determinant = 0

Neonatal AND mortalities AND determinants = 0

Neonatal AND mortalities AND outcome = 0

Neonatal AND mortalities AND outcomes = 0

Supplementary Tables

Supplementary Table 1: Neonatal mortality rates per 1,000 live births in the European Region, 2000–2021. Mean neonatal mortality rates (NMR) per 1,000 live births (LB) for 28 countries in the European Region. Data are based on WHO estimates. The linear regression coefficient (Lm coeff) represents the annual rate of change in neonatal mortality over time. SD: standard deviation.

| Country | NMR/1,000 LB | | Percentage Change | Lm coeff | P value |
| --- | --- | --- | --- | --- | --- |
|  | **mean** | **sd** |  |  |  |
| Austria | 2.53 | 0.32 | -31.48% | -0.050 | <0.001 |
| Belgium | 2.45 | 0.22 | -24.69% | -0.031 | <0.001 |
| Cyprus | 2.17 | 0.57 | -45.26% | -0.076 | <0.001 |
| Czechia | 1.87 | 0.37 | -48.04% | -0.054 | <0.001 |
| Denmark | 2.93 | 0.30 | -39.47% | -0.038 | <0.001 |
| Estonia | 2.41 | 1.32 | -83.39% | -0.200 | <0.001 |
| Finland | 1.73 | 0.38 | -45.34% | -0.057 | <0.001 |
| France | 2.48 | 0.13 | -6.77% | -0.005 | 0.264 |
| Germany | 2.43 | 0.18 | -19.20% | -0.027 | <0.001 |
| Greece | 2.59 | 0.45 | -39.03% | -0.039 | 0.007 |
| Hungary | 3.48 | 1.06 | -61.91% | -0.163 | <0.001 |
| Iceland | 1.48 | 0.23 | -36.00% | -0.030 | <0.001 |
| Ireland | 2.72 | 0.56 | -47.41% | -0.080 | <0.001 |
| Italy | 2.41 | 0.48 | -52.26% | -0.073 | <0.001 |
| Latvia | 4.07 | 1.59 | -74.94% | -0.248 | <0.001 |
| Lithuania | 3.08 | 0.93 | -55.47% | -0.141 | <0.001 |
| Luxembourg | 1.70 | 0.21 | -25.14% | -0.018 | 0.006 |
| Malta | 4.46 | 0.28 | -23.77% | -0.040 | <0.001 |
| Netherlands | 3.00 | 0.44 | -32.15% | -0.064 | <0.001 |
| Norway | 1.91 | 0.42 | -50.46% | -0.065 | <0.001 |
| Poland | 3.76 | 0.98 | -53.10% | -0.150 | <0.001 |
| Portugal | 2.26 | 0.43 | -48.91% | -0.059 | <0.001 |
| Slovakia | 3.52 | 0.62 | -39.98% | -0.088 | <0.001 |
| Slovenia | 1.97 | 0.60 | -58.39% | -0.090 | <0.001 |
| Spain | 2.18 | 0.32 | -36.60% | -0.048 | <0.001 |
| Sweden | 1.73 | 0.28 | -38.67% | -0.041 | <0.001 |
| Switzerland | 3.11 | 0.20 | -17.16% | -0.032 | <0.001 |
| United Kingdom | 3.12 | 0.35 | -26.18% | -0.052 | <0.001 |
| All countries | 2.63 | 0.96 | -44.5% | -0.074 | <0.001 |

Supplementary Table 2: Study characteristics for all the included studies

|  | **Title** | **First Author, year and country** | **Published at** | **Design** | **Population** | **Singleton/**  **Multiple** | **Results (relevant to this systematic review)** | **Is data from an open-access database?** | **Grade-score – NOS** | **Suitable for meta-analysis?** |
| --- | --- | --- | --- | --- | --- | --- | --- | --- | --- | --- |
| 1 | **Neonatal organ donation: Retrospective audit into potential donation in a single neonatal unit (1)** | Ali et al.  UK, London  2023 | Wiley – Nursing in critical care | Audit  **Assessed as a Cross-sectional study for NOS.** | All neonatal deaths at a NICU in London  2012-2021. | Not available | 189 neonatal deaths on the neonatal unit between January 2012 and December 2021.  Causes of neonatal death (number, %):   - *Complications arising from prematurity (70, 37%) - Congenital malformations or abnormalities (63, 33%) - Perinatal asphyxia (37, 20%) - Miscellaneous (18, 9.5%) - Sepsis (1, 0.5%)   *Complications arising from prematurity included: low gestational age, respiratory distress syndrome, patent ductus arteriosus, bronchopulmonary dysplasia, intraventricular haemorrhage and necrotizing enterocolitis. | No | Selection: ***  Comparability:  Outcome: *  Total stars: 4 | No, only have data for at term neonates. |
| **2** | **Application of ICD-PM to preterm-related neonatal deaths in South Africa and United Kingdom (2)** | Allanson et al.  UK  2016 | Wiley – BJOG – An international Journal of Obstetrics and Gynaecology | Retrospective application of ICD-PM  **Assessed as a Cross-sectional study for NOS.** | The UK database with all perinatal deaths (*n* = 9067) in the West Midlands  1997 - 2010. | Both singleton and multiple births are included. | Study population: 956 term neonatal deaths and 3248 preterm neonatal deaths.  Neonatal death causes are divided in all, preterm and at term.  Causes of all neonatal deaths (ICD-PM classification, %):   - Low birthweight and prematurity (N9, 31.6%) - congenital malformations, deformations, and chromosomal abnormalities (N1, 27.4%) - Neonatal deaths of unspecified cause (N11, 26.1%) - Respiratory and cardiovascular disorders (N7, 6.8%) - Other neonatal conditions (N8, 2.5%) - Complications of intrapartum events (N4, 1.7%) - Infection (N6, 1.5%) - Miscellaneous (N10, 1.3%) - Convulsions and disorders of cerebral status (N5, 0.6%) - Birth trauma (N3, 0.3%) - Disorders related fetal growth (N2, 0.3%) | No | Selection: ***  Comparability:  Outcome: *  Total stars: 4 | Yes |
| **3** | **Causes of death among full term stillbirths and early neonatal deaths in the Region of Southern Denmark (3)** | Basu et al.  Denmark, Region South.  2017 | J. Perinat. Med. | **Cross-sectional study** | Early neonatal deaths within 7 days of life in the Region of Southern Denmark  January 1, 2010 – December 31, 2014. | Not available | 95 maternal-infant cases were included, of which 21 of the deaths were neonatal.  Only neonatal death within the first 7 days of life.  Causes of death according to the CODAC classification system (number, %):   - Congenital anomaly (6, 28.6%) - Unknown (6, 28.6%) - Neonatal (5, 23.8%) - Intrapartum (2, 9.5%) - Placenta (1, 4.8%) - Maternal (1, 4.8%) - Infection (0, 0) - Fetal (0, 0) - Cord (0, 0)   Death causes according to the INCODE-based classification system (number, %):   - Fetal genetic, structural and karyotypic abnormalities (7, 33.3%) - Unknown (6, 28.6%) - 3Other pertinent condition not specified (4, 19.0%) - Obstetric complications (3, 14.3%) - Maternal or fetal hematological conditions (1, 4.8%) | No | Selection: ***  Comparability:  Outcome: *  Total stars: 4 | No, only have data for at term neonates. |
| **4** | **Assessing the deprivation gap in stillbirths and neonatal deaths by cause of death: a national population-based study (4)** | Best et al.  UK (England, Wales, Scotland and the UK Crown Dependencies)  2019 | BMJ - Journals.  Arch Dis Child Fetal Neonatal | Retrospective cohort study  **Assessed as a Cross-sectional study for NOS.** | Singleton births.  1 January 2014 - 31 December 2015  ≥24 weeks’ gestation. | Singleton | Study population: 2345 neonatal deaths.  Causes of neonatal death (number, %):   - Congenital anomaly (774, 33%) - 24-27 weeks not SGA (475, 20%) - Other/unknown (236, 10%) - Neurological (203, 8.7%) - 28-31 weeks not SGA (185, 7.9%) - 37+ weeks SGA (105, 4.5%) - Cardiorespiratory (75, 3.2%) - Infection (73, 3.1%) - 24-27 weeks SGA (63, 2.7%) - Fetal (45, 1.9%) - 28-31 weeks SGA (41, 1.8%) - Intrapartum (38, 1.6%) - 32-36 weeks SGA (32, 1.4%) | No | Selection: ***  Comparability: **  Outcome: **  Total stars: 7 | Yes |
| **5** | **Circumstances, causes and timing of death in extremely preterm infants admitted to NICU: The EPIPAGE-2 study (5)** | Boileau et al.  France  2023 | Wiley – Acta pædiatrica | **Cohort study.** | Preterm infants.  Born GA 22 - 34 weeks.  France  2011  The EPIPAGE-2 population-based cohort. | Singleton and multiple births are included. | Neonatal deaths:   - 0-2 days: 53 - 3-7 days: 61 - 8-28 days: 79   % of death causes are read from a figure, therefor approximately.  Data is not publicly available.  “All deaths” also contains infants > 28 days, therefor this category is not used.  Death causes 0-2 days: (app. %)   - Respiratory disease (55%) - Unknown (15%) - Other (15%) - Infection (10%) - CNS injury (5%)   Death causes 2-7 days: (app. %)   - CNS injury (49%) - Respiratory disease (38%) - Other (8%) - Infection (5%)   Death causes 8-28 days: (app. %)   - CNS injury (35%) - Respiratory disease (28%) - Infection (20%) - Other (6%) - Unknown (6%) - Necrotizing enterocolitis (5%) | No | Selection: ****  Comparability: *  Outcome: ***  Total stars: 8 | No |
| **6** | **[Causes of perinatal deaths in children delivered out of hospital in material collected by Chair and Department of Forensic Medicine, Medical University of Warsaw] (6)**  ***The article is in polish.*** | Borowska-Solonynko et al.  Poland  2011 | Arch Med Sadowej Kryminol | **Cross-sectional study** | 27 cases of out of hospital born fetuses and newborns.  2001-2008.  Department of Forensic Medicine at the Medical University of Warsaw | Both singleton and multiple are included. | At risk group: out of hospital births.  Total of neonatal deaths: 10  Death causes established through autopsies.  Neonatal death causes: (number)   - Prematurity (3) - Injuries resulting from fetal head maladaptation (3) - Multiorgan trauma (2) - Violent suffocation (1) - Unknown (1) | No | Selection: *  Comparability:  Outcome: *  Total stars: 2 | No, forensic autopsies. |
| **7** | **Neonatal mortality in NHS maternity units by timing and mode of birth: a retrospective linked cohort study (7)** | Carty et al.  England  2023 | BMJ - Journals | **Retrospective cohort** linking birth registration, birth notification and hospital episode data | 6.054.536 liveborn singleton births.  2005 – 2014  NHS maternity units in England.  Liveborn with GA < 22 weeks are excluded. | singleton | 13,077 Neonatal deaths.  Wigglesworth cause of death categories are used to modify the neonatal deaths.  Death causes (number):  Cause arises before the onset of labour:   - Immaturity-related conditions (n = 6,178) - Congenital anomalies (n = 4,070) - Antepartum infections (n = 489)   Cause arises during, or shortly after labour and birth:   - Asphyxia, anoxia or trauma (n = 1,494)   Cause arises after birth:   - Infections (n = 237) - Sudden infant deaths (n = 190) - Other specific conditions (n = 111) - External conditions (n = 44)   Unclassified:   - Other conditions (n = 239) - Missing (n = 25) | No | Selection: ****  Comparability: *  Outcome: ***  Total stars: 8 | Yes |
| **8** | **Diagnosis and cause of death in a neonatal intensive care unit--how important is autopsy? (8)** | Costa et al.  Portugal  2011 | The Journal of Maternal-Fetal & Neonatal Medicine | Retrospective review of NICU autopsies  **Assessed as a Cross-sectional study for NOS.** | 110 deaths in the NICU of a hospital in Porto from 2004-2008.  Median GA = 34 weeks  Median death age: 10,7 days.  Age at death 0-317 days | Not available | 53 neonatal patients with autopsies.  Causes of death divided in subgroups. In Class 1A and 1b the cause for each neonate is represented.  Class 1A (clinical diagnosis – Pathology diagnosis):   - Meningitis with septic shock – Hirschsprung disease, peritonitis and sepsis; no meningitis - Primary pulmonary hypertension; persistence of fetal circulation - Thrombosis of right pulmonary artery and branches of renal vein - Post-operatory of aortic coarctation - Luminal thrombus with total occlusion of the prosthesis   Class 1B: (clinical diagnosis – Pathology diagnosis):   - Renal insufficiency and hydropsia. probable nephrotic syndrome - Congenital hemochromatosis: no signs of nephrotic syndrome - Malformative syndrome (not identified) - Campomelic dysplasia - Meconial ileus; cystic fibrosis mutation negative - Changes highly suggestive of cystic fibrosis - Hepatic insufficiency (unknown etiology) - Congenital Hemochromatosis - Malformative syndrome (not identified) - Malformative syndrome related to cloacal persistence with pulmonary hypoplasia (due to compression)   Class II (n = 20):   - infection, most frequently pneumonia in patients already receiving antibiotics, but also one case of enterovirus infection - vascular accidents such as myocardial infarction and intestinal ischemia - congenital malformations namely two cases of noncompac- tion cardiomiopathy with tricuspid valve anomaly - hemophagocytic syndrome ( n = 1) - small-bowel perforation by dialysis catheter with peritonitis in a patient in palliative care due to congenital nephrotic syndrome (n = 1)   Class III (n = 2)   - central nervous system hemorrhage (n = 1) - Pulmonary hypoplasia (n = 1)   Class IV (n = 5)   - congenital malformations - leukomalacia - Negative findings (pulmonary hypoplasia, hyaline membrane disease, among other diagnoses, that were not confirmed by autopsy)   Class V (n =18) | No | Selection: ***  Comparability:  Outcome: *  Total stars: 4 | No, because of the way the data is represented |
| **9** | **Prevention of early-onset Group B Streptococcal disease - the Northern Ireland experience (9)** | Eastwood et al.  Northern Ireland,  2015 | BJOG | Retrospective observational study.  **Assessed as a Cross-sectional study for NOS.** | 43 neonates with EOGBS out of a sample of 779 cases. | Not available | At risk population: Early onset GBS  5 neonates with EOGBS died.  Causes:   - Sepsis (n = 3) - Other (n = 2) | No | Selection: ***  Comparability:  Outcome: *  Total stars: 4 | No, too specific at-risk group. |
| **10** | **Trends in cause and place of death for children in Portugal (a European country with no Paediatric palliative care) during 1987-2011: a population-based study (10)** | Forjaz de Lacerda et al.  Portugal  2017 | BMC Pediatr | **Cross-sectional** epidemiological population- based study | 38,870 child deaths in Portugal from 1987 to 2011.  Death causes were divided into three categories: CCS, other medical causes and trauma.  10,571 were caused by CCS (complex chronical conditions). | Not available | Risk factor population: CCS (complex chronical conditions).  3055 neonatal deaths with CCS from 1987 to 2011.  CCC categories (numbers):   - Cancer (20) - Neuromuscular (449) - Cardiovascular (1005) - Respiratory (193) - Renal (135) - Gastrointestinal (88) - Haem. And immun. (0) - Metabolic (38) - Other (1127) | No | Selection: ***  Comparability: **  Outcome: **  Total stars: 7 | No, too specific at-risk group |
| **11** | **Intentional child and adolescent homicides in Milan (Italy): A 30-year interdisciplinary study (11)** | Galante et al.  Italy, Milan  2024 | Legal Medicine - Elsevier | Retrospective study on cases of child and adolescent homicides.  **Assessed as a Cross-sectional study for NOS.** | 28,302 autopsy reports in Milan from January 1991 to December 2020. | Not available | Risk factor population: Intentional homicides.  7 Neonaticides  Death causes:   - Drowning (n = 3) - Multiple sharp injuries (n = 2) - Traumatic brain injury (n = 1) - Starvation and hypothermia (n = 1) | No | Selection: ***  Comparability:  Outcome: **  Total stars: 5 | No, forensic autopsies. |
| **12** | **Contribution of Congenital Anomalies to Neonatal Mortality Rates in Malta (12)** | Gatt et al.  Malta  2015 | Paediatric and Perinatal Epidemiology | **Cross-sectional study** | 84 821 livebirths with 441 neonatal deaths.  1994–2013  Malta. | Not available | Total neonatal deaths 1994-2013: 441  Attributed to congenital anomaly: 162  Attributed to non-congenital perinatal causes: 279 | No | Selection: ***  Comparability:  Outcome: *  Total stars: 4 | No, congenital anomalies |
| **13** | **Inequalities in mortality of infants under one year of age according to foetal causes and maternal age in rural and urban areas in Poland, 2004-2013 (13)** | Genowska et al.  Poland  2016 | Annals of Agricultural and Environmental medicine | **Cross-sectional study** | Polish population 2004-2013.  Information from the Central Statistical Office. | Not available | Neonatal mortality rates by cause rural areas: (median, %)   - All-causes of NMR (3.93, 100) - Perinatal conditions (mainly due to a preterm birth, intrauterine asphyxia and infections of the perinatal period) (2.5, 62.6) - Congenital malformations (1.2, 32.6) - SIDS (0.03, 0.9) - Infectious and parasitic diseases 0.03 - External causes 0.03 - Respiratory system 0.02   Neonatal mortality rates by cause urban areas: (median, %)   - All-causes of NMR 3.99 - Perinatal conditions (mainly due to a preterm birth, intrauterine asphyxia and infections of the perinatal period) (2.68, 66.4) - Congenital malformations (1.06, 28.8) - SIDS (0.04, 1.0) - Infectious and parasitic diseases 0.05 - External causes 0.02 - Respiratory system 0.02 | No | Selection: ***  Comparability:**  Outcome: **  Total stars: 7 | Yes |
| **14** | **Abandonment of newborn infants: a Danish forensic medical survey 1997-2008 (14)** | Gheorghe et al.  Denmark,  2011 | Forensic Sci Med Pathol | Forensic medical survey  **Assessed as a Cross-sectional study for NOS.** | 12 newborn infant corpses found abandon. 1997-2008. | Not available | Risk factor population: Corpses found abandont  Death causes based on autopsies.  Five were alive at birth and had the following death causes:   - Anoxia neonatorum (n = 1) - Suffocation (n = 2) - Asphyxia (n = 1) - Brain injury (n = 1) | No | Selection: ***  Comparability:  Outcome: *  Total stars: 4 | No, forensic autopsies. |
| **15** | **Stillbirth and neonatal mortality in monochorionic and dichorionic twins: a population-based study (15)** | Glinianaia et al.  UK, North of England  2011 | Hum Reprod | Population-based study  **Cohort study** | 9130 twins born in North of England between 1998 and 2007 | Monochorionic and dichorionic twins | Risk factor population: twins  201 neonatal mortalities.  Causes of death (number):   - Prematurity* (129) - Infection (26) - Congenital anomaly (22) - Intrapartum hypoxia or trauma (10) - Twin-twin transfusion syndrome (9) - Other miscellaneous perinatal illness (3) - Antepartum hypoxia (1) - SIDS (1)   * Include: severe pulmonary immaturity, hyaline membrane disease and intraventricular hemorrhage. | No | Selection: ****  Comparability: *  Outcome: ***  Total stars: 8 | No, twins |
| **16** | **Cause of intrauterine and neonatal death in twin pregnancies (CoDiT): development of a novel classification system (16)** | Gulati et al.  UK, West midlands  2020 | BJOG | Retrospective **cross-sectional study** | Twin pregnancies in the West Midlands  1 January 2005 and 31 December 2016 | Monochorionic (MC) and dichorionic (DC) twins | Risk factor population: twins  A total of 16 neonatal deaths.  Neonatal death causes (number):   - Placental (6) - Prematurity (6) - Congenital abnormality (2) - Other (1) - Unknown (1) | No | Selection: ***  Comparability: *  Outcome: **  Total stars: 6 | No, twins |
| **17** | **Survival analysis of a cohort of extremely preterm infants born in Finland during 2005-2013 (17)** | Harkin et al.  Finland,  2021 | J Matern Fetal Neonatal Med | **Cohort study** | Extremely preterm infants born in Finland during 2005-2013 | Both singleton and multiple are included. | Risk factor population: extremely preterm  Obs. > 7 days death  Neonatal death causes:  Mortality during the first 48 h was mainly due to extreme immaturity regardless of intrauterine growth.  During 48–168 neonatal hours (from 2 to 7 days) SGA:   - RDS (36.8% of all deaths) - Pulmonary hemorrhage (31.6%) - Pulmonary hypertension (10.5%)   Non-SGA:   - Severe IVH (58% of all deaths) - Respiratory deaths (16% of all deaths).   SGA > 7 days of life:   - Severe BPD (31.8%) - NEC (13.6%) - severe IVH (13.6%), - RDS (9.1%).   Non-SGA:   - NEC (32.4%), - severe IVH (17.6%), - sepsis (14.7%) - severe BPD (11.8%).   SGA > 2 days of life:   - Pulmonary causes (60.9%) - Non-respiratory deaths (39.1%)   Non-SGA   - Pulmonary causes (29.8%) - Non-respiratory deaths (70.2%) | No | Selection: ****  Comparability: *  Outcome: ***  Total stars:8 | Yes, preterm |
| **18** | **Histological findings in unclassified sudden infant death, including sudden infant death syndrome (18)** | Liebrechts-Akkerman et al.  The Netherlands  2013 | Pediatr Dev Pathol | A multicenter retrospective study  **Assessed as a Cross-sectional study for NOS.** | 200 Dutch unclassified sudden infant death cases.  1984 - 2005. | Not available | Risk factor population: unclassified sudden infant death  Perinatal: (n = 3)   - 1 day old: Lungs – bronchopneumonia - 1 day old: Lungs – Massive aspiration - 1 day old: Lungs and brain – Bronchopneumonia and meningitis   SIDS A: (n =1)   - 3 weeks old: kidney and thyroid - Calcifications and fetal remnants, respectively   SIDS B: (n = 1)   - 2 weeks old: blood culture – pneumococcus   < 1 year B: (n = 1 )   - 2 weeks old: lungs - Staphylococcus aureus in swab | No | Selection: ***  Comparability:  Outcome: *  Total stars: 4 | No, can be used as an example for the future regarding autopsies and classification of death causes. |
| **19** | **Global, regional, and national causes of under-5 mortality in 2000-15: an updated systematic analysis with implications for the Sustainable Development Goals (19)**  **OBS DATA WILL BE TAKEN FROM THE DATABASE** | Liu et al.  Global,  2016 | Lancet | An update to the annual estimates of child mortality by cause to 2000–2015  **Assessed as a Cross-sectional study for NOS.** | 2.7 million neonatal deaths. | Not available | Neonatal death causes by country 2015.  PNE = pneumonia  PRE = preterm  INT = Intrapartum related events  SEP = Sepsis/meningitis  OTH = Other disorders  CON = Congenital  In total: 11328 (number, %)   - PNE (87, 0.77%) - PRE (4604, 40.64%) - INT (1383, 12.21%) - SEP (579, 5.11%) - OTH (1425, 12.58%) - CON (3341, 29.49%) | Yes | Selection: ***  Comparability:  Outcome: *  Total stars: 4 | No, since it uses data from a database. |
| **20** | **Child homicide and neglect in France: 1991-2008 (20)** | Makhlouf et al.  France  2014 | Child Abuse Negl | Retrospectively investigation of autopsies  **Assessed as a Cross-sectional study for NOS.** | Child homicide and death by neglect at the age of 15 or less in France between 1991 and 2008. | Not available | Risk factor population: homicide / death by neglect  Total neonatal deaths: 16  Causes of neonatal death (number, %):   - Neglect (8, 50%) - Asphyxia/drowning (5, 31.2%) - Head trauma (3, 18.8%) | No | Selection: ***  Comparability:  Outcome: *  Total stars: 4 | No, forensic autopsies |
| **21** | **[The Importance of Autopsy in Early Neonatal Death in Portugal] (21)**    **Article is in Portuguese** | Miranda et al.  Portugal  2020 | Acta Med Port | Retrospective study of clinical records.  **Assessed as a Cross-sectional study for NOS.** | All neonates who died during the first week of life in a NICU in Portugal.  2008 – 2017 | Both singleton and multiple are included. | 76 neonatal deaths within the first week of life.  Wigglesworths classification for neonatal death causes: (number, %)  Congenital anomalies (28, 36.8%)   - CNS disorder (2) - Cardiovascular disorder (6) - Digestive disorders (1) - Urinary system disorders (2) - Musculoskeletal disorders (6) - Chromosomal anomalies (1) - Non-chromosomal polymalformative syndrome (3) - Others (7)   Immaturity or preterm birth (33, 43.4%)   - Hyaline membrane disease (12) - Infection (7) - Necrotizing enterocolitis (4) - CNS haemorrhage (9) - Others (1)   Intrapartum asphyxia (11, 14.5%)   - Placental abruption (4) - Others (7)   Other specific causes (4, 5.3%)   - CNS disorder (1) - Cardiovascular disorder (1) - Metabolic disorder (1) - Maternal bacterial infection (1) | No | Selection: ***  Comparability: *  Outcome: **  Total stars: 6 | Yes |
| **22** | **Cause-Specific Stillbirth and Neonatal Death According to Prepregnancy Obesity and Early Gestational Weight Gain: A Study in the Danish National Birth Cohort (22)** | Nohr et al.  Denmark  2021 | Nutrients | Prospective cohort study  **Cohort study** | 85,822 pregnancies in the Danish National Birth Cohort (1996–2002) | Singleton | 226 neonatal deaths  Causes of death: (number, %)   - Congenital anomalies (91, 40.3%) - Placental dysfunction (15, 6.6%) - Umbilical cord complications (2, 0.9%) - Maternal disease (12, 5.3%) - Intrapartum events (26, 11.5%) - Preterm birth (32, 14.2%) - Infections (23, 10.2%) - Other causes* (25, 11.1%)   * Other causes contained other specific conditions (23) and unclassifiable (10) | No | Selection: ****  Comparability: **  Outcome: ***  Total stars: 9 | Yes |
| **23** | **Retrospective analysis of neonatal deaths secondary to infections in England and Wales, 2013-2015 (23)** | Oligbu et al.  England and Wales  2021 | Arch Dis Child Fetal Neonatal Ed | Retrospective analysis of national electronic death registrations data  **Assessed as a Cross-sectional study for NOS.** | England and Wales.  2013–2015.  2,091,597 neonates. | Not available | 5095 neonatal deaths from 2013-2015 in England and Wales  GA >22 weeks  Neonates aged <28 days.  **669 infection-related deaths.**  Divided further into: Term, preterm, extremely preterm and all.  Also divided further in detail into different kinds of pathogens. | No | Selection: ***  Comparability:  Outcome: *  Total stars: 4 | No, infections |
| **24** | **Perinatal and neonatal mortality in Northwest Greece (1996-2004) (24)** | Pantou et al.  Greece  2010 | J Matern Fetal Neonatal Med | Analysis of birth and death registers  **Assessed as a Cross-sectional study for NOS.** | 25,960 births in NW Greece in the period 1996-2004. | Not available | 103 Neonatal deaths.  GA > 22 weeks or BW > 500 g  Death causes: (number)   - Respiratory Distress Syndrome (36) - Sepsis (17) - Infections (5) - Congenital infections (2) - Congenital anomalies (11) - Hypoxic ischaemic encephalopathy (4) - Periintraventricular haemorrhage (5) - Extreme prematurity (6) - Hyperkalemia (1) - Kernicterus (1) - Hydrops foetalis (1) - Aspiration pneumonia (1) - Necrotising enterocolitis (2)   Early neonatal death (0-7 days) causes are also described separately (n = 41). | No | Selection: ***  Comparability: *  Outcome: *  Total stars: 5 | Yes |
| **25** | **Pathogen-specific mortality in very low birth weight infants with primary bloodstream infection (25)** | Piening et al.  Germany  2017 | PLoS One | **Cohort study** | 55,465 very low birth weight infants.  The German national neonatal infection surveillance system NEO-KISS | Not available | At risk population: very low birth weight infants  Neonatal deaths: 4094 VLBW infants with MCBSI (microbiology confirmed blood stream infections) | No | Selection: ****  Comparability: **  Outcome: ***  Total stars: 9 | No, low birthweight |
| **26** | **[The analysis of neonatal deaths based on autopsy protocols of the Department of Forensic Medicine in Bialystok in the years 1955-2009] (26)**  **Article in polish** | Ptaszynska-Sarosiek et al.  Poland,  2011 | Arch Med Sadowej Kryminol | Post-mortem examination of autopsies  **Assessed as a Cross-sectional study for NOS.** | 17,838 forensic medical autopsies.  Department of Forensic Medicine in Białystok  1955-2009 | Not available | At risk population: forensic  124 cases of neonatal death.  Older newborns: 16  Causes of death older newborns (number):   - Pneumonia (3) - Gastroenteritis (1) - Homicide / accident (8) - Failure to assist (2) - Undetermined (2)   Causes of death newborns (a few days old) (number):   - Suspicion of active neonaticide (56) - Suspicion of passive neonaticide (36) - Disease (2) - Childbirth trauma (4) - Undetermined (10) | No | Selection: ***  Comparability:  Outcome: *  Total stars: 4 | No, forensic |
| **27** | **High mortality among children with gastroschisis after the neonatal period: A long-term follow-up study (27)** | Risby et al.  Denmark,  2017 | J Pediatr Surg | Long-term follow up study  **Cohort study** | 71 infants referred to primary surgery for gastroschisis defect.  January 1, 1997, to December 31 2009. | Not available | At risk population: children with gastroschisis  4 neonatal deaths:   - Congenital Cytomegalo-virus-infection with intracerebral and intestinal bleeding (n = 1) - Cardiac arrest, pericardial tamponade with PN due to displacement of CVK (n = 1) - Septicaemia induced multiorgan failure (n = 1) - Acute small bowel obstruction (n = 1) | No | Selection: ****  Comparability:  Outcome: *  Total stars: 5 | No, too specific at-risk group |
| **28** | **Causes of death in children with congenital anomalies up to age 10 in eight European countries (28)** | Rissmann et al.  Denmark, Finland, Italy, Malta, Netherlands, Norway, Spain and the UK  Regions differ in each country. EUROCAT based.  2023 | BMJ Paediatr Open | Register based cohort study  **Assessed as a Cross-sectional study for NOS.** | 7386 dead children with congenital anomalies | Not available | At risk population: congenital anomalies  Children with congenital anomalies born alive between 1 January 1995 and 31 December 2014.  4199 neonatal deaths.  Underlying causes of death for neonates with CA:   - CA 71% - Infections 1% - Immaturity 7% - Other 11% - Missing 9%   The underlying cause of death for each CA is described. | No | Selection: ***  Comparability:  Outcome: **  Total stars: 5 | No, congenital anomalies |
| **29** | **Examination of (suspected) neonaticides in Germany: a critical report on a comparative study (29)** | Schulte et al.  Germany  2013 | Int J Legal Med | Report on a comparative study  **Assessed as a Cross-sectional study for NOS.** | Files of 150 deceased children in Germany from 1993-2007 | Not available | At risk population: suspected neonaticides.  Autopsies  Cause of death for 57 neonates (number):   - All forms of suffocation (41) - Head injury (8) - Still birth / intrauterine hypoxia (5) - Drowning (5) - Neglect (3) - Hypovolemic shock (4) - Hypothermia (2) - Intracranial birth injury (1) - Fetal asphyxia during delivery (1) - Combined causes of death (6) | No | Selection: ***  Comparability:  Outcome: *  Total stars: 4 | No, forensic |
| **30** | **Socioeconomic inequalities in outcome of pregnancy and neonatal mortality associated with congenital anomalies: population based study (30)** | Smith et al.  East Midlands and South Yorkshire regions of England  2011 | BMJ | Retrospective population-based registry study  **Assessed as a Cross-sectional study for NOS.** | All registered cases of nine selected congenital anomalies with an end of pregnancy date between 1 January 1998 and 31 December 2007. | Not available | At risk population: congenital anomalies  Neonatal deaths by congenital anomalies adjusted for year of birth:  1,98 /10.000 live births.  They also have an adjusted for maternal age estimate. | No | Selection: ***  Comparability: **  Outcome: **  Total stars: 7 | No, congenital anomalies |
| **31** | **Nature of socioeconomic inequalities in neonatal mortality: population based study (31)** | Smith et al.  England  2010 | BMJ | Retrospective cohort study.  **Assessed as a Cross-sectional study for NOS.** | 18 524 neonatal deaths.  Singletons born between 1 January 1997 and 31 December 2007. | Singleton | Causes of neonatal death (number, %):   - Congenital anomalies (4464, 24.1%) - Intrapartum events (1959, 10.6%) - Immaturity < 24 weeks’ gestation (3602, 19.4%) - Immaturity 24-27 weeks’ gestation (3603, 19.5%) - Immaturity 28-36 weeks’ gestation (916, 4.9%) - Immaturity (gestation unknown) (117, 0.6%) - Infection (1677, 9.1%) - Accident and other specific causes (1295, 7.0%) - Sudden infant death (543, 2.9%) - Unclassified (348, 1.9%) | Yes, GBD. | Selection: ***  Comparability:  Outcome: **  Total stars: 5 | No, will use the GBD database |
| **32** | **Neonatal Morbidity and 1-Year Survival of Extremely Preterm Infants (32)** | Stensvold et al.  Norway  2017 | Pediatrics | Prospective population-based study  **Cohort study** | All infants born at 22 through 26 weeks’ gestation in Norway in 2013–2014 | Both singleton and multiple are included. | At risk group: extremely preterm infants (GA 22-27 weeks)  NICU death (n=66) causes: (number, %)   - Respiratory failure (22, 33.3%) - Severe neurologic morbidity (19, 28.8%) - NEC or bowel perforation (13, 19.7%) - Sepsis (5, 7.6%) - Congenital malformation (1, 1.5%) - Perinatal asphyxia (1, 1.5%) - Accidental extubation (1, 1.5%) - Refractory hypotension (4, 6.1%) | No | Selection: ****  Comparability: *  Outcome: ***  Total stars: 8 | Yes, preterm |
| **33** | **Major contributors to hospital mortality in very-low-birth-weight infants: data of the birth year 2010 cohort of the German Neonatal Network (33)** | Stichtenoth et al.  Germany  2012 | Klin Padiatr | Prospective **cohort study** | 2221 VLBW infants born in 46 NICUS in the German Neonatal Network from January until December 2010. | Both singleton and multiple are included. | At risk group: very-low-birth-weight infants.  Major contributors to mortality: (*approximately number)   - RDS (43) - BPD (5) - Pulmonary haemorrhage (11) - Other respiratory failure (6) - Sepsis (37) - NEC/FIP (19) - IVH grade 4 (24) - Comfort care (25) - Lethal malformations (14) - Others (26) - Not determined (11)   *Obs. Numbers are read from a figure. | No | Selection: ****  Comparability: **  Outcome: ***  Total stars: 9 | No, low birthweight |
| **34** | **Why do young children die in the UK? A comparison with Sweden (34)** | Tambe et al.  The UK and Sweden  2015 | Arch Dis Child | Comparative study  **Assessed as a Cross-sectional study for NOS.** | Total of 14104 deaths aged < 5 years in the UK.  1036 deaths aged < 5 years in Sweden.  Period: 2006-2008 | Not available | Neonatal deaths in the period:   - UK 8960 - Sweden 574   ICD-codes are used for death causes.  Mortality rates 0-27 days UK: (rates pr. 100.000)   - P0-96 Certain conditions originating in the perinatal period (296.9) - Q0-99 Congenital malformations and chromosomal anomalies (68.6) - R0-99 Symptoms, signs, abnormal clinical and laboratory findings not elsewhere classified (7.8) - G0-99 Diseases of the nervous system (4.0) - V0-Y89 External causes of morbidity and mortality (3.7) - A0-B99 Certain infectious and parasitic diseases (0.4) - J0-J99 Diseases of the respiratory system (0.1) - C0-D48 Neoplasms (0.8) - I0-99 Diseases of the circulatory system (2.7) - E0-90 Endocrine, nutritional and metabolic diseases (4.1) - K0-93 Diseases of the digestive system (0.7) - D50-89 Diseases of the blood and blood forming organs and immune system (0.4) - L0-99 Diseases of the skin and subcutaneous tissue (0.1)   Mortality rates 0-27 days Sweden: (rates pr. 100.000)   - P0-96 Certain conditions originating in the perinatal period (97.2) - Q0-99 Congenital malformations and chromosomal anomalies (50.0) - R0-99 Symptoms, signs, abnormal clinical and laboratory findings not elsewhere classified (21.0) - G0-99 Diseases of the nervous system (2.5) - V0-Y89 External causes of morbidity and mortality (1.3) - A0-B99 Certain infectious and parasitic diseases (3.5) - J0-J99 Diseases of the respiratory system (0.3) - C0-D48 Neoplasms (1.6) - I0-99 Diseases of the circulatory system (1.3) - E0-90 Endocrine, nutritional and metabolic diseases (1.9) - K0-93 Diseases of the digestive system (0.9) - D50-89 Diseases of the blood and blood forming organs and immune system (0.3) | No | Selection: ***  Comparability:  Outcome: **  Total stars: 5 | Yes. |
| **35** | **[Causes of stillbirths and perinatal death in Poland between 2007-2009] (35)**  **Article in polish** | Troszynski et al.  Poland  2011 | Ginekol Pol | Cohort study  **Assessed as a Cross-sectional study for NOS.** | Women who gave birth in 2007-2009 from 12 provinces.  N = 614,816 | Not available | *** Google imagetranslater was used.  Death up to 7 days of age (n: 1412). ICD is used for causes. (number, %)   - Q00-Q99 Developmental defects (435, 30.8) - P05-P08 Disorders related to the duration of pregnancy and fetal growth (500, 35.4) - P00-P91 Other causes (294, 20.8) - P20-P28 Respiratory disorders (183, 13.0) | No | Selection: ***  Comparability:  Outcome: *  Total stars: 4 | Yes |
| **36** | **Survival and causes of death in extremely preterm infants in the Netherlands (36)** | van Beek et al.  The Netherlands  2021 | Arch Dis Child Fetal Neonatal Ed | National **cohort study** | 3312 stillborn and live born infants.  GA 24^0/7^ - 26^6/7^ weeks  Born between January 2011 and December 2017  Data from the Netherlands Perinatal Registry. | Both singleton and multiple are included. | Extremely preterm infants  Causes of death is divided in GA 24. GA 25, GA 26 and total.  Total = 603 neonates.  The article uses the classification by Patel *et al* to divide causes of death.  Causes of death for all the involved neonates (total group): (number, %)   - NEC (168, 27.9%) - RDS (115, 19.1%) - Severe intracranial haemorrhage (100, 16.6%) - Infection (96, 15.9%) - BPD (65, 10.8%) - Other (41, 6.8%) - Congenital malformation (8, 1.3%) - Immaturity (5, 0.8%) - Non-classifiable (5, 0.8%) | No | Selection: ****  Comparability: *  Outcome: ***  Total stars: 8 | Yes, preterm |
| **37** | **Perinatal complications in twin pregnancies after 34 weeks: effects of gestational age at delivery and chorionicity (37)** | Vergani et al.  Italy  2013 | Am J Perinatol | **Cross-sectional study** | A cohort of 471 twin pregnancies with GA > 34. | Twins | At risk population: twins  5 neonatal deaths – causes:   - IVH, DIC - DIC, RDS - PDA, IVH, DIC, RDS - RDS - Sepsis, RDS | No | Selection: *  Comparability:  Outcome: *  Total stars: 2 | No, twins |
| **38** | **Categorizing neonatal deaths: a cross-cultural study in the United States, Canada, and The Netherlands (38)** | Verhagen et al.  The US, Canada and the Netherlands  2010 | J Pediatr | **Cross-sectional study** | Newborn > 22 weeks GA, who died in the delivery room or the NICU.  October 2005 to September 2006  4 NICUs (Chicago, Milwaukee, Montreal, and Groningen) | Not available | 52 NICU deaths in the Netherlands (Groningen)  Causes of death: (number, %)   - Asphyxia (9, 17%) - Congenital anomalies (26, 50%) - Sepsis/NEC (12, 23%) - Respiratory insufficiency (1, 2%) - Intracranial bleeding (4, 8%) | No | Selection: ***  Comparability: *  Outcome: **  Total stars: 6 | Yes |
| **39** | **The changing profile of infant mortality from bacterial, viral and fungal infection over two decades (39)** | Williams et al.  Northern England  2013 | Acta Paediatr | Population-based survey  **Assessed as a Cross-sectional study for NOS.** | 4366 infant deaths from a population of 704 536 livebirths.  1988 - 2008  North England | Not available | Infection was the cause of death for 13% of infant deaths in the population.  Total early neonatal death by infection: 213  Total late neonatal death by infection: 127  Early neonatal death causes (number):   - Bacteria (142) - Virus (8) - Fungi (2) - Unknown (61)   Late neonatal death causes (number):   - Bacteria (88) - Virus (14) - Fungi (8) - Unknown (18) | No | Selection: ***  Comparability: **  Outcome: **  Total stars: 7 | No, infections |
| **40** | **Viral infections: contributions to late fetal death, stillbirth, and infant death (40)**  **** same population as the article above **** | Williams et al.  Northern England  2013 | J Pediatr | Population-based survey  **Assessed as a Cross-sectional study for NOS.** | 4366 infant deaths from a population of 704 536 livebirths.  1988 - 2008  North England | Not available | 2796 neonatal deaths. 340 of these were caused by infection.  Viral Infection contributed to 6.5% of neonatal deaths = 22. | No | Selection: ***  Comparability:**  Outcome: *  Total stars: 6 | No, infections.  The same data as the article above. |
| Additional articles found through included articles | | | | | | | | | | |
| **41** | **Short term outcomes after extreme preterm birth in England: comparison of two birth cohorts in 1995 and 2006 (the EPICure studies)**  **(41)** | Costeloe et al.  2012  England | BMJ | **Cross-sectional study** | 2006:  3133 born 22-26 GA weeks.  1115 born 22-25 GA weeks.  1995:  666 born 22- 25 weeks GA. | Both singleton and multiple are included and accounted for. | 400 NICU deaths in January to December 2006.  Causes of death: (number, %)   - Congenital malformation (4, 1) - Pulmonary insufficiency (85, 16) - Respiratory distress syndrome / intracerebral haemorrhage / infection (174, 33) - Late sequelae of ventilation (40, 8) - Infection (84, 16) - Intracranial haemorrhage (33, 6) - Necrotising enterocolitis (63, 12) - Other (35, 7) - Not known (4, 1) | No | Selection: ***  Comparability:**  Outcome: **  Total stars: 7 | Yes, preterm |

### Supplementary Figures

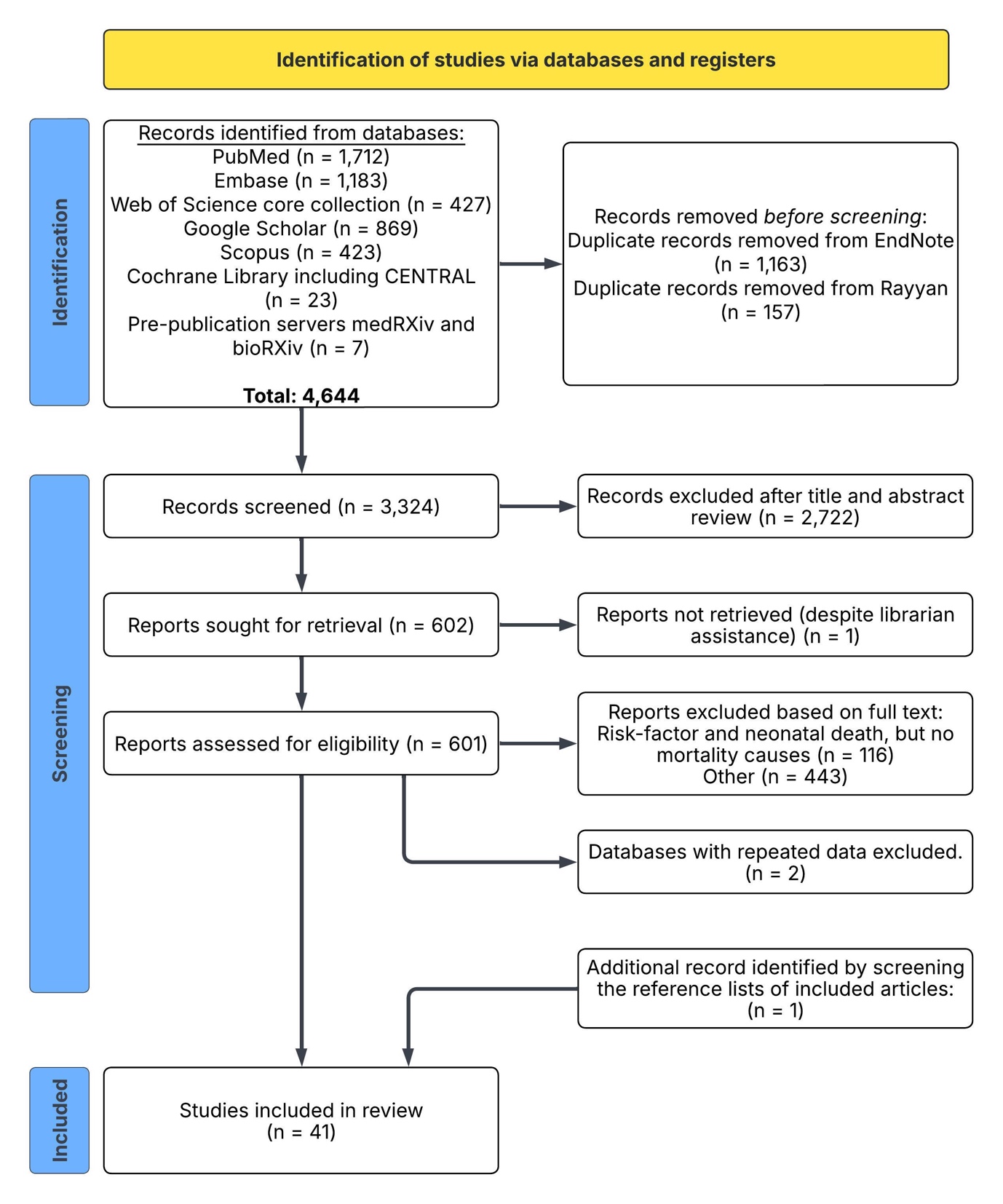

Supplementary Figure 1: Flowchart of the article selection process. PRISMA flow diagram illustration the process of study identification, screening, eligibility assessment, and inclusion for the systematic review.

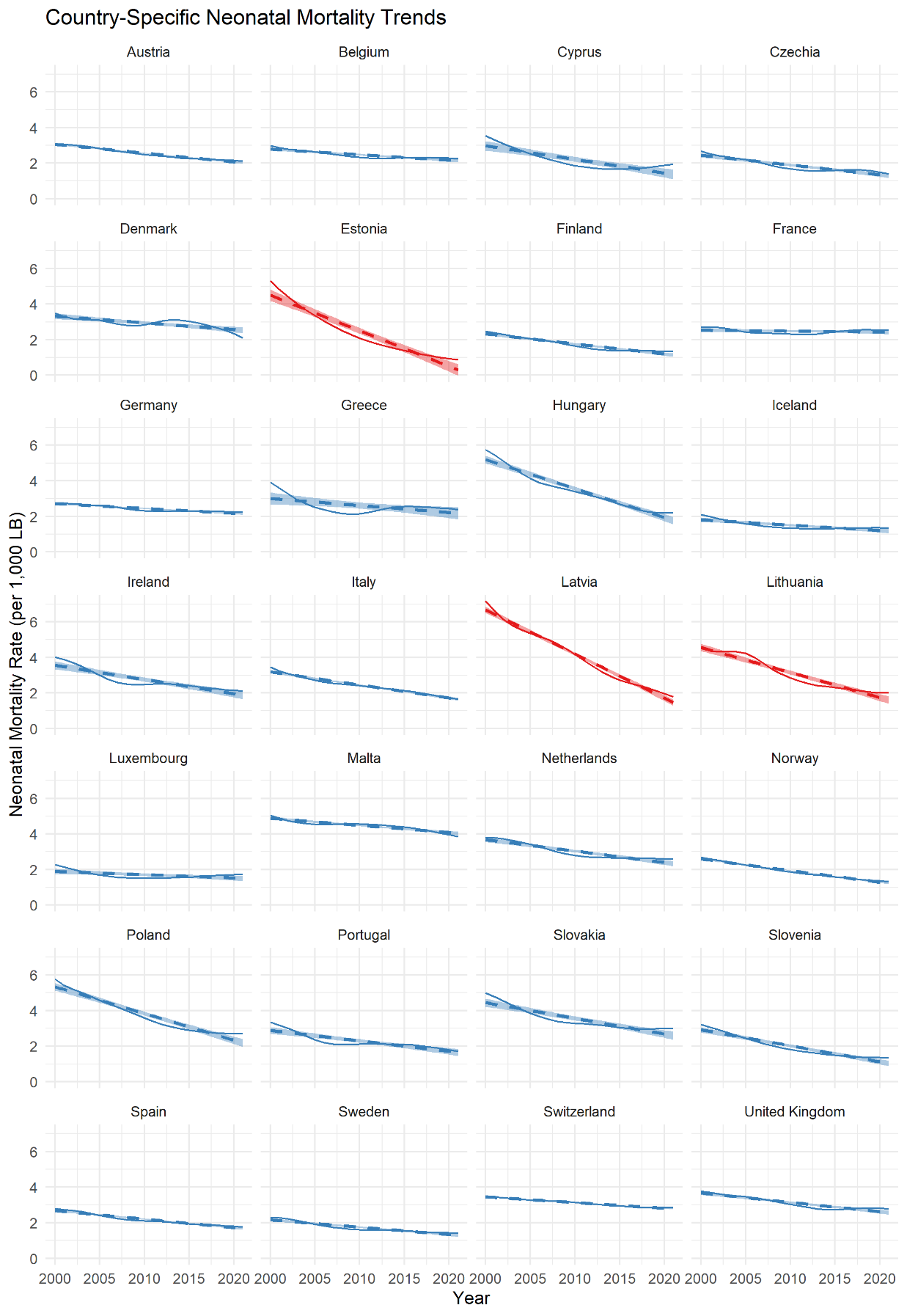

Supplementary Figure 2: Country-specific Neonatal Mortality Trends, 2000–2021. The solid lines represent the annual neonatal mortality rate per 1,000 live births for each included country. The shaded dashed lines represent a linear regression model fitted to the data. The regression coefficient (Coef.) indicate the decline in neonatal mortality over time. Colours reflect clusters identified within the data; Estonia, Latvia, and Lithuania (Red) represent a small cluster which is distinct from the other countries (Blue)
